# Development of a National Stroke Audit to Enhance the Quality of Acute Stroke Care in Ireland: A Scoping Review and Delphi Consensus Synthesis

**DOI:** 10.1101/2024.10.08.24313597

**Authors:** Catherine Nora Moran, Isabelle Jeffares, Joan McCormack, Niamh A. Merriman, Carlos Bruen, Agnes Jonsson, Paul J. Murphy, Khyber Afridi Rabbi, Joseph Harbison, David J Williams, Peter J. Kelly, Rónán Collins, Eithne Sexton, Frances Horgan, Máirín Ní Bheacáin, Elaine Byrne, John Thornton, Collette Tully, Anne Hickey

## Abstract

**Background:** Population ageing, treatment advances, evolving models of care, and between-hospital heterogeneity in patient outcomes underscore the need for continual audit to ensure the delivery of high-quality, equitable, and evidence-based stroke care. This study aimed to develop a core minimum dataset for acute stroke care in Ireland, for integration into the Irish National Audit of Stroke (INAS), benchmarked against, and aligned with, international best practice.

**Methods:** This scoping review was conducted in accordance with the Preferred Reporting Items for Systematic Reviews and Meta-Analysis Extension for Scoping Reviews (PRISMA-ScR). Medline Ovid, Embase, CINAHL EBSCOhost, and pertinent grey literature were searched from 2010 to identify national and continuous stroke audits. Titles, abstracts, and full texts were assessed for eligibility. Audit documentation was retrieved from identified eligible audits and stroke care data items were extracted, translated, and charted. Data charting enabled comparison of existing Irish audit items with frequently collected international items to identify commonalities and/or gaps in coverage. Acute stroke care items were then reviewed by key stakeholders in a three-round Delphi consultation.

**Results:** Twenty-one eligible international stroke audits and registries were identified, containing ∼4,500 audit items. Key stakeholders were consulted for their expert perspectives on the existing Irish (*n*=103), frequently collected international (*n*=97), and additional expert-suggested (*n*=22) acute items regarding their potential inclusion in INAS. Following consensus, a core minimum dataset comprising 86 acute care and 35 thrombectomy items was finalised.

**Conclusions:** The findings of this scoping review represent the “ideal” core outcomes dataset for acute stroke care in Ireland, derived from international benchmarking and stakeholder consultation. This dataset serves as the “gold standard” for monitoring acute stroke care in Ireland, aimed at enhancing patient outcomes, and supporting local and national quality improvement initiatives.

## Introduction

Stroke is the second most common single cause of mortality in the European Union (EU) (1) and the predominant reason for acute hospitalisations in high-income countries (2-4). It is a leading contributor to adult-acquired disability globally (4, 5), leading to a range of short-term to persistent post-stroke sequelae including functional, communication, psychological, and cognitive deficits (6-9). Additionally, stroke poses a considerable societal and economic burden with respect to direct and indirect (e.g., informal care and productivity loss) healthcare costs (10-14). The cost of stroke is estimated at €45 billion within Europe (1) and accounts for 4% of the Irish annual healthcare expenditure (15).

In Ireland, approximately 6,000 newly diagnosed stroke cases are admitted to hospital annually (15, 16), with an estimated 30,000-45,000 stroke survivors living in the community or long-term care (12); a figure predicted to double by 2035 (17). In the EU, there are approximately 1.1 million incident strokes and over 9.5 million stroke survivors, the prevalence of which is projected to increase by 27% by 2047 (13). Owing to significant improvements in acute stroke management (18, 19) and advances in primary and secondary prevention strategies, the age-specific rates of new strokes and stroke mortality have decreased over the last two decades, corresponding with improved patient survival and outcomes (16, 20, 21). However, given the incremental demographic growth of the ageing population (22), the absolute incidence of stroke is rising (23). As such, the number of stroke survivors living with chronic disability and specialist care needs, and consequent burden on families, healthcare systems, and expenditure, is increasing and will likely keep increasing in line with age profile projections (13, 24, 25).

Variability exists in stroke outcomes reported across hospitals (16, 26-28). Such heterogeneity may arise from differing availabilities of post-stroke therapies or varying care quality, highlighting the need for ongoing and continual review of stroke healthcare services to ensure high-quality and equitable best practice is being delivered. By prospectively and continuously tracking consecutive stroke admissions, national clinical audit makes it possible to record the proportion of patients receiving care that meets clinical standards and discern any variation in quality or access across hospitals, regions, or time periods (16). Stroke audits also allow for the identification of specific care processes and treatments that are associated with better patient outcomes (29, 30). As such, audit data can be used to monitor trends and benchmark against international standards in a way that can be used by healthcare decision-makers to institute policy change, identify care priorities, and address service delivery gaps. Indeed, countries with hospital-based national clinical audits or registries of stroke care with routine data collection have seen corresponding improvements in stroke health care, patient outcomes, and changes in health policy (31, 32).

A national stroke register for Ireland was developed originally in 2010-2011. In 2019, governance of the register was taken over by the National Office of Clinical Audit (NOCA) (33) and became the Irish National Audit of Stroke (INAS) (34). INAS is a clinically-governed continuous national quality audit with a web-based portal recording data on in-patients with acute stroke from hospital sites across Ireland. Key performance indicators are measured against Irish (35, 36) and United Kingdom (UK) (37) guidelines, and joint UK-Ireland guidelines (38). INAS sources demographic, clinical, and administrative data from an existing national database of discharges from acute public hospitals in Ireland (Hospital In-Patient Enquiry (HIPE) system). It collects stroke-specific clinical data on all patients who have experienced a stroke (‘Core Clinical’ dataset), submitted from each hospital to HIPE via a stroke audit portal. Additionally, thrombectomy data (‘Thrombectomy’ dataset) are collected on patients who receive a thrombectomy from the two Endovascular Thrombectomy (EVT) centres in Ireland. Since 2018, separate rehabilitation and recovery data are collected by health and social care professionals (HSCPs) in participating hospitals (‘HSCP’ dataset).

The aim of this research was to further develop INAS by establishing a core minimum dataset for acute care (Phase 1), based on a scoping review of international practice and created collaboratively through iterative cycles of stakeholder engagement. The development of the audit dataset for rehabilitation and recovery care (Phase 2) and the identification of resourcing needs and production of an implementation strategy (Phase 3) will be reported separately.

## Methods

### Protocol and registration

The protocol for this scoping review was published (39) and registered with OSF Registries (40). The scoping review was conducted in accordance with the six-stage stepwise methodological framework specified by Arksey and O’Malley and advanced by Levac et al. and Peters et al. (41-43). The resultant scoping review findings are reported in line with the Joanna Briggs Institute (JBI) Preferred Reporting Items for Systematic Reviews and Meta-Analysis Extension for Scoping Reviews (PRISMA-ScR) reporting guidelines (44).

### Eligibility criteria

Peer-reviewed and grey literature were systematically searched for national and continuous international stroke audits and registries published since 2010. In line with the recommended PCC (population, concept, context) approach, our inclusion criteria comprised:

- **Population:** Stroke (any type).
- **Concept:** National stroke audit, defined as a data collection program (register, databank, or database) used for measuring and monitoring the quality and structure of stroke care services and performance indicators across multiple participating sites for patients hospitalised with stroke (45).
- **Context:** The stroke audit or registry operated as the established national country-wide system for stroke care data collection, bore the country’s name (as guided by the names of the United Nations member states, or constituent country of a member state, and including Taiwan), or included the term ‘national’ within its title. Audits pertaining to care quality across both acute and non-acute stroke care settings were considered for inclusion. Only audits and registries with continuous data collection, that had at least one year of prospective data collection, and that were still functioning in 2021 were included.

### Information sources

Electronic databases (Medline Ovid, Embase, and CINAHL EBSCOhost) were searched for peer-reviewed literature published in English since 2010 to identify national stroke audits or registries. A targeted search for relevant grey literature was also performed by searching Google Scholar and websites of interest, such as stroke organisation websites and reports (e.g., the European Stroke Organisation (46), NHS National Stroke Service Model 2021 (47), etc.). A broad range of published and unpublished evidence sources were eligible, including primary research studies, systematic reviews, meta-analyses, website reports, guidelines, and recommendations. Conference abstracts were excluded. This search strategy was developed in consultation with an information specialist (PM) and is consistent with the search conducted within a similar systematic review (45).

### Search

Medline Ovid, Embase, and CINAHL EBSCOhost were searched from 2010 using medical subject headings (MeSH) and text words on the 10th of February 2021, and then updated on the 4^th^ of February 2024. Search terms included “stroke”, “transient ischaemic attack”, “intracerebral haemorrhage”, “national stroke registry”, “stroke register”, and “stroke audit” (**Supplemental File A**). All results were imported into EndNote X9.3.3 and duplicates were removed. To ensure literature saturation, a snowball approach was adopted; as sources of evidence were identified, the reference lists (ascendency search) and papers that cited the sources (descendency search) were scanned for relevant materials. Once an eligible audit was identified, a more specific search approach was employed to obtain information regarding audit datasets, data dictionaries, and data collection procedures. Where the documentation was not published or available on the audit website, authors/audit managers were contacted. Audit documents that were provided in a language other than English were translated and included in the data charting and synthesis stages.

### Selection of sources of evidence

Two independent reviewers from a bank of reviewers (CB, NM, KR, GH, AH) initially screened titles and abstracts against pre-defined inclusion and exclusion criteria (39). Following this, the full texts for potentially relevant sources were retrieved and further assessed for eligibility. At each stage, discrepancies were resolved through consensus-based discussion and consultation with a third reviewer (AH), when necessary. The current Irish stroke audit (INAS) was considered within the scope of the review and was included in the charting and synthesis stages to ensure that the Irish audit was reviewed alongside international practice.

### Data charting process

The previous scoping review stages allowed for the inclusion of sources of evidence relating to national stroke audits of all types. During the data charting and synthesis steps, the included documents were categorised as relating to acute (*Phase 1*) or non-acute (*Phase 2*) stroke care (Phase 2 is reported separately).

Data relating to stroke care audits were extracted by two independent researchers (CNM, NM) from the retrieved documentation and charted into a Microsoft Excel proforma. A data charting calibration check was conducted whereby data extracted by both reviewers for a random sample of one third of the databases were compared. The level of inter-rater agreement was 95%, and any discrepancies (e.g., missing data or uncertainty) were resolved through consensus-based discussion. As agreement was high, data charting for the remaining audit documents was conducted by either one of the researchers. The results of the data charting exercise were discussed with the INAS Governance Committee in an iterative process that facilitated, where appropriate, the potential revision of the charting framework to ensure that data were captured comprehensively. Any missing details were sought through additional searching or further contact with the relevant audit report authors.

### Data items

The following stroke care items were extracted from the international stroke audit documentation: (a) audit characteristics and context (setting, phase of care covered, eligible patient population, number of sites, follow-up) and (b) acute audit data items (audit question/item, response options).

### Critical appraisal of individual sources of evidence

The quality of the individual sources of evidence was not formally assessed as the aim of a scoping review is to provide an overview of the breadth and current status of available evidence on a topic from different source types and not to assess the weight or robustness of the evidence (41, 43).

### Synthesis of results

#### International Benchmarking of Acute Stroke Care Items

Once the data items were collated, a two-stage benchmarking exercise was conducted whereby the acute stroke care items present across the retrieved international audit databases were cross-checked to identify commonalities and/or gaps in coverage. In the *first* stage, we identified which items the Irish stroke audit shared with international stroke audits and registries. With INAS as the key comparator, each of the Irish Core Clinical and Thrombectomy items was cross-checked against all other included international audits and registries to look for equivalent or closely related acute stroke care items. Frequency counts, representing the number of audits that contained the same (or closely related) item, were tallied for each INAS item to show which Irish items are commonly or infrequently collected internationally, or perhaps are unique to the Irish healthcare context.

In the *second* stage, we aimed to identify which other acute items were collected internationally (and how frequently) that were not captured by INAS. With the Australian register (AuSCR (48)) as the key comparator (chosen as it represented the most comprehensive stroke registry), any additional acute stroke care items were cross-checked to see how frequently other international audits and registries asked something equivalent or closely related. After cross-checking the INAS and AuSCR audits against all other included international audits, any additional items found in remaining international audits were cross-checked. Frequency counts were generated to display how many international audits collected each acute data item that the Irish audit does not currently collect.

The benchmarking process generated an inventory of acute stroke care items currently in use (Irish audit and international items) with frequency mapping detailing how common or uncommon each item is in international practice. Following consultation with the project Steering Group, a criterion was set such that only predominant international items, that is, those items shared amongst four or more audits/registries in total (i.e., the comparator audit item and at least 3 other audits/registries), were put forward for further consideration by a stakeholder panel (i.e., *n*=97 international items).

#### Stakeholder Delphi Consultation on the Acute Stroke Care Dataset

Stakeholders were invited to participate in a multi-round Delphi consultation to review existing INAS items (*n*=103) and the inventory of the most frequently collected international acute stroke care items (*n*=97) (49, 50). The Delphi panel included 24 participating members^a^ of the INAS Governance Committee and project Steering Group, comprising stroke and health services researchers (*n*=4), patient representatives (*n*=2), personnel working in national stroke policy, patient safety, and service planning (*n*=5), stroke clinicians (*n*=8), and allied healthcare professionals with stroke expertise (*N*=5: *n*=1 physiotherapy^b^; *n*=3 nursing; *n*=1 psychology).

Across three rounds of the Delphi consultation, stakeholders provided expert opinions on all of the existing Irish audit items, independent of the benchmarking frequencies, and evaluated whether the current INAS Core Clinical and Thrombectomy items should be retained in, or omitted from, INAS. The panel were also asked to consider whether any of the frequently collected international items should be added to INAS as part of the “ideal” core minimum acute stroke care outcome dataset. Additionally, in the third round, stakeholders reviewed 22 new items suggested in previous voting rounds and derived from stakeholder discussions. For Rounds 1 and 2, participants voted to “Include in INAS”, “Exclude from INAS”, “No strong view”, or “Don’t know enough to be able to say”, and provided feedback. For the final round, participants voted according to a reduced set of responses: “Include in INAS”, “Exclude from INAS”, or “Don’t know enough to be able to say”. Stakeholders were advised to base their decisions on the importance of the data for the Irish audit, disregarding any current logistical and resource limitations.

Following Round 1 of the process, participants could revise their initial responses after reviewing anonymised responses and feedback from other group members. This iterative process aimed to build consensus while minimising the potential undue influence of group dynamics. The results from each Delphi round were categorised according to consensus criteria based on the mean percentage of votes to include the item in INAS. Specifically, Irish, international, and additional expert-suggested acute stroke care items were each classified as receiving high (≥70%), moderate (50-69%), or low (<50%) support. A consensus threshold of ≥70% for inclusion of the item as part of the ideal acute stroke care outcome dataset for INAS was set *a priori* to the task administration. Detailed voting instructions for each Delphi round are provided in **Supplemental File B**.

## Results

### Selection of Sources of Evidence

The initial scoping review search yielded 5,396 records, of which 263 duplicates were removed. Two independent reviewers initially screened 5,133 titles and abstracts for eligibility. Subsequently, 871 full texts were reviewed, from which 27 national stroke audits and registries were identified, including the Irish stroke audit. After further review, four of the identified stroke audits/registries were later excluded owing to non-continuous data collection at the time of the search, namely, the Chinese National Stroke Registry (51), Finnish National Stroke Database (52), Malaysian National Neurology Registry (53), and the Thai Stroke Registry (54). An additional two stroke registries were excluded as we were unable to obtain the relevant audit documentation or information detailing the data items, despite numerous requests (the Polish Stroke Registry (55, 56) and the Russian National Stroke Registry (57)). Included audits comprised the Australian Stroke Clinical Registry (AuSCR) (48), the Austrian Stroke Unit Registry (58), CorHealth Ontario (Canada) (59), DanStroke (Denmark) (60, 61), the German Stroke Register Study Group (ADSR) (62-64), the Irish National Audit of Stroke (INAS) (34), the Japan Stroke Data Bank (65-67), the Dutch Acute Stroke Audit (Netherlands) (68), the Norwegian Stroke Registry (NHR) (69, 70), the Scottish Stroke Care Audit (SSCA) (71), the Singapore Stroke Registry (72), the Slovak Stroke Register (73), the Clinical Research Collaboration for Stroke in South Korea (CRCS-K) (74), the Korean Stroke Registry (KSR; South Korea) (75), RENISEN (Spain) (76), Riksstroke (Sweden) (31, 77), the Swiss Stroke Registry (78, 79), the Taiwan Stroke Registry (80), the Sentinel Stroke National Audit Programme (SSNAP; UK) (81, 82), Get with the Guidelines Stroke Registry (GWTG; USA) (83), and the Paul Coverdell National Acute Stroke Programme (PCNASP; USA) (84). Audit documentation for five of the included stroke registries were available in languages other than English and were translated (58, 61, 64, 67, 69).

The database search was updated in February 2024, resulting in 1591 additional titles since the initial search. Following removal of 418 duplicates, 1173 titles and abstracts were screened for eligibility. Following exclusions, 755 records were retrieved for review. Only one additional national stroke audit/registry was identified: the Israeli National Stroke Registry^c^ (85, 86). After numerous requests, we were unable to obtain the necessary registry documentation, leading to its exclusion from the review.

As follows, twenty-one stroke audits/registries (across 19 countries) with national coverage and continuous data collection were included in the scoping review (see **Figure 1** flow diagram).

[**Figure 1**]

### Characteristics of Sources of Evidence

This review incorporates 21 national and continuous audits and registries, the characteristics of which are detailed in **Supplemental File C**. Notably, both the United States (83, 84) and South Korea (74, 75) each hosted two national stroke registries. Most of the audits and registries initiated data collection in the late 1990s and early-to-mid 2000s and all remain operational. All audits and registries collected data on patients with ischaemic and haemorrhagic stroke, and two-thirds of registries also included patients with a transient ischaemic attack (TIA). Audits and registries varied with respect to whether there was voluntary involvement by participating hospitals or whether data acquisition was government mandated.

### Results of Individual Sources of Evidence

#### Results of the International Benchmarking of Acute Stroke Care Items

Following data extraction, an inventory of the acute stroke care data items from 21 included stroke audits and registries was compiled. This inventory comprised almost 4,500 items derived from 20 international stroke audits/registries, and a further 103 items from the Irish stroke audit. Specifically, the Irish audit included 10 items from an administrative database (HIPE), 58 items comprising the ‘Core Clinical’ dataset, and 35 items comprising the ‘Thrombectomy’ dataset.

During the initial phase of data charting, the Irish acute audit items were systematically compared with those from the 20 other international audits and registries. The frequency counts, depicted in **Supplemental File D**, illustrate how many international audits or registries collect similar (or closely related) items for each of the INAS acute care items. The tabulated frequency counts excluded the INAS index item from the tally. In brief, 15, 48, and 40 INAS items were shared by a low, moderate, or high number of eligible international registries, respectively.

The second data charting stage involved comparing the coverage of all acute stroke care items not captured by INAS across each of the 20 international stroke audits/registries. Items that appeared in four or more international stroke audits/registries in total (i.e., the comparison audit item and three or more other audits/registries) were selected for review in the subsequent stakeholder consultation. A total of 97 such frequently occurring acute stroke care items were identified. **Supplemental File E** presents frequency counts depicting how many international audits incorporate each of the 97 acute stroke care data items that are not currently encompassed in INAS. Among these international items, 90 were utilised by a moderate number of stroke registries, while 7 were shared by a high number of registries.

### Synthesis of Results

#### Results of the Stakeholder Delphi Consultation

##### Delphi Round 1

Twenty out of 24 eligible stakeholders responded to Round 1 of the Delphi exercise (83% response rate). The group voting averages for each response option pertaining to the INAS and international audit items are presented in **Supplemental Files F and G**, respectively. After Round 1, 56/103 INAS items received a high percentage of votes (≥70%) for inclusion (or in this case, retention) as part of INAS (specifically, 9/10 HIPE, 42/58 Core Clinical, and 5/35 Thrombectomy items) (**Supplemental File F**). Of the 97 frequently collected international items, 17 items obtained high votes (≥70%) for inclusion in (or in this case, addition to) INAS (**Supplemental File G**).

##### Delphi Round 2

In Round 2 of the consultation, the initial group of responding stakeholders (*n*=20) reviewed their previous responses alongside the mean aggregated group responses for each audit item under consideration. Of these stakeholders, 11 altered their responses for several items. On average, responses were changed for 4.9% of the items. A further 6.6% of the votes cast during Round 2 addressed items that had missing data in Round 1. Eight stakeholders did not respond to the opportunity to change their previous responses, which was interpreted as no change. One additional stakeholder left their position after Round 1, hence, their previous responses also remained unchanged. The Round 2 percentages displayed in **Supplemental Files F and G** represent the combined percentage votes across Rounds 1 and 2 for the 20 stakeholders, including the initial ‘unchanged’ votes for the 9 stakeholders who did not indicate any changes in Round 2, and the amended votes for the 11 stakeholders who did.

Following Round 2, 61 INAS items (i.e., 10/10 HIPE, 44/58 Core Clinical, and 7/35 Thrombectomy items) garnered a high percentage of votes (≥70%) for inclusion (or retention) in INAS (**Supplemental File F**). Eleven Core Clinical items and 20 Thrombectomy items received a moderate percentage of votes (50-69%) and only 3 Core Clinical and 8 Thrombectomy items received a low proportion of votes (<50%) for inclusion. Among the 97 international items, 24 items obtained a high percentage of votes for inclusion as part of INAS, 25 received moderate votes and 48 received a low proportion of votes (**Supplemental File G**). None of the items from the inventory of Irish or international stroke audit items were highly voted for exclusion.

Based on these results, the low-voted INAS Core Clinical (*n*=3) and low-voted international (*n*=48) items were excluded from further consideration. The highly-voted Irish items (*N*=61) were recommended for ongoing retention in INAS without the need for further deliberation. An additional 22 items, derived from stakeholder input, were subsequently added to the inventory of audit items for consideration in Round 3, alongside the remaining items that required consensus (i.e., the low-voted INAS Thrombectomy items, the INAS Core Clinical and international items with moderate endorsement, and the highly-voted international items).

##### Delphi Round 3

In the third Round, 17/22 eligible stakeholders (77% response rate) voted on, and discussed in a subsequent stakeholder meeting, the remaining inventory of items to finalise their potential inclusion in the ideal core dataset. Two INAS Core Clinical items received a high percentage of votes for inclusion and hence were recommended for ongoing retention in INAS (**Supplemental File F**). Six Core Clinical items received a moderate proportion of votes for inclusion, while 3 Core Clinical items received a low proportion of votes; these items did not meet the inclusion cut-off and were, therefore, suggested for exclusion from the final dataset. Regarding the remaining INAS Thrombectomy items, 6 items were moderately-voted and 22 were low-voted. According to our inclusion cut-off, these items should be excluded. However, following consultation with the stakeholders, a decision was made that the Thrombectomy dataset should be retained in its entirety to support the development of a National Thrombectomy Service in Ireland.

For the remaining items under consideration, 24 international items and 6/22 additional expert-suggested items were highly-voted and thus, were proposed for inclusion in INAS (**Supplemental File G**). Sixteen moderately-voted and 9-low voted international items, along with 10 moderately-voted and 6 low-voted additional expert-suggested items, were excluded from INAS.

#### Final “Ideal” Core Minimum Dataset for Acute Stroke Care in Ireland

Following the international benchmarking and multi-round stakeholder consultation processes, a consensus was achieved on the final ideal core minimum dataset for acute stroke care in Ireland, totalling 121 items (**Table 1**). The final ideal dataset retains 10 items from HIPE, 46 Core Clinical items, and 35 Thrombectomy items. Additionally, it incorporates 24 international items and 6 expert-suggested items. The recommended additions primarily examine history of known stroke-specific risk factors, complications during hospital admission, details on medications before admission and at discharge, screening for aphasia and cognitive impairment, and some further acute clinical details. The Thrombectomy dataset is collected exclusively at two EVT centres in Ireland. Therefore, the final recommended dataset for capturing acute clinical data across *all* participating hospitals comprises 86 items.

[**Table 1**]

## Discussion

This paper details the development of a core minimum acute stroke care dataset for integration into the Irish National Audit of Stroke (INAS), designed in accordance with international best practice as identified through a scoping review of international stroke audits and refined through iterative cycles of stakeholder engagement.

We identified 24 national, continuous, hospital-based clinical stroke registries and audits, spanning 22 countries (including Ireland), that are currently in use to monitor the quality of acute stroke care. Three of these audits did not respond to repeated contact from the research team requesting their audit items (55-57, 85, 86); thus, 21 audits were included in this scoping review analysis. The final consensus-derived ideal dataset for Ireland comprises 121 items in total: 35 items specific to thrombectomy collected at two EVT centres only, and 86 acute clinical items for national data collection. Despite numerous thrombectomy items meeting the criterion for exclusion, stakeholder discussions led to the decision to retain the Thrombectomy dataset in its entirety to support the ongoing development of the National Thrombectomy Service in Ireland. Regarding the 86 acute clinical items, 56 were retained from the original INAS Core Clinical dataset, while 30 new items – a combination of international items and items identified by the stakeholder panel - were recommended for inclusion to address identified gaps in data capture. Twelve items from the original dataset did not meet the cut-off for inclusion for the final ideal dataset, suggesting potential redundancy and/or the need for revision. The proposed changes, namely those items recommended for addition to, or elimination from, INAS are discussed in the following sections.

### Additional items recommended for inclusion in INAS

A number of items recommended for inclusion in INAS require alterations to current national IT infrastructure. For example, Ireland does not currently have a unique patient identifier system, albeit legislation was published in July 2024 providing the legal basis for establishing individual digital health records (87). If implemented, this would facilitate valuable linkage of patient data across hospitals and care pathways, and enable follow up into the community. Such data linkage is critical to the success of many other international stroke audits (31, 77, 81, 82).

Data relating to arrival to hospital by ambulance and the documentation of stroke severity were recommended for inclusion in INAS. Both items are commonly collected by international audits. Recording NIHSS data was identified by stakeholders as vital for better understanding of stroke severity nationwide, enabling more equitable delivery of appropriate acute treatments and more precise mortality predictions.

The largest grouping of new items recommended for inclusion in INAS were 12 items pertaining to history of known stroke risk factors. Risk factor items are widely collected in stroke audits internationally; for example, history of ‘*diabetes*’ (*n*=17 international audits/registries), ‘*previous stroke’* (*n*=15), ‘*hypertension*’ (*n*=15), ‘*previous TIA*’ (*n*=13), ‘*hypercholesterolaemia*’ (*n*=9), etc. Information on risk factor history is crucial for effective stroke prevention and awareness campaigns. It also plays a vital role in tracking changes in risk factor profiles as population demographics shift, facilitating the identification of vulnerable groups, and the adjustment of strategies to improve survival outcomes.

Increasing the number of items relating to pre- and post-stroke medication use was also recommended. At present, INAS includes items that record antithrombotic therapy prior to admission (for those with known atrial fibrillation), during acute treatment, and at discharge (as secondary prevention). Following data synthesis, three additional medication items were recommended for inclusion in INAS, relating to anti-hypertensive use and lipid-lowering treatment prior to this stroke (collected by *n*=8 and *n*=6 international audits/registries, respectively), and prescription of anti-hypertensive agents on discharge from stroke. These therapies were identified as crucial for reducing the risk of ischaemic and recurrent strokes by targeting risk factors such as ‘hypertension’ (88) and ‘hypercholesterolaemia’ (89).

Some audit items from international audits were recommended by stakeholders for inclusion in INAS as periodic and/or local audits rather than continuous national collection. Items relating to hospital-based complications of stroke, such as ‘*aspiration pneumonia*’ (collected by *n*=7 international audits/registries), ‘*urinary tract infection*’ (*n*=5), and ‘*seizures*’ (*n*=5), were identified as valuable for revealing care areas needing intervention, highlighting service disparities, and informing service improvements.

Finally, six items did not meet the initial inclusion cut-off from the international benchmarking exercise but were later suggested for inclusion during stakeholder discussions and were highly voted in Round 3 of the Delphi consultation. The ‘*location of intracerebral hematomas in intracranial haemorrhage (ICH)’* was deemed worthy of inclusion in INAS as different aetiologies may influence prevention strategies and such proportional data are lacking. Stakeholders also recommended including the ‘*modified Rankin Scale (mRS) at 90 days*’, to supplement existing INAS items that capture the mRS pre-stroke and at discharge. Incorporating a 90-day outcome score aligns with international standards, is vital for tracking patient outcomes, comparing performance and monitoring service trends, and for adjusting metrics by cohort severity.

Stakeholders recommended that two items be moved from the INAS HSCP dataset to the Core Clinical dataset, specifically ‘*was screening for cognitive impairment completed using a valid screening measure?*’ and ‘*does the patient have aphasia?*’, and for these to be supplemented with ‘*was screening for aphasia conducted?*’ and ‘*diagnosis of pre-stroke dementia*’. Although clinical guidelines highlight the need for cognitive screening (38, 90), its implementation is inconsistent owing to ongoing challenges surrounding the need for valid stroke-specific screening tools (90), limited access to clinical neuropsychologists for stroke (16), uncertainty regarding the optimal time for assessments, and the fluctuating nature of post-stroke cognitive difficulties during and following an acute hospital stay. Nevertheless, cognitive screening is a vital first step, alongside more comprehensive neuropsychological and functional assessments, for identifying deficits and determining immediate support needs, and longer-term rehabilitation planning (90).

### Existing items recommended for exclusion from INAS

Twelve existing INAS Core Clinical items were recommended for exclusion. A number were secondary items, providing free-text spaces to specify reasons not listed in a previous item’s response set. Free text was considered to be effortful to input, difficult to ensure data quality, and challenging to analyse. The recommendation is that item response sets be as comprehensive as possible, eliminating the need for follow up free text items.

Other items were deemed not to be meaningful (e.g., “*case complete*”) and therefore, were recommended for exclusion. Finally, some items were recommended for exclusion from INAS in their current format. Specifically, three items referring to carotid stenosis were recommended for exclusion and/or consideration for item revision. As an important risk factor for stroke, stakeholders proposed that the phrasing of international audit items relating to carotid stenosis be considered in rephrasing the Irish audit items in this domain. Stakeholders also recommended that further item iterations could benefit from specifying whether carotid imaging was performed initially, the severity of stenosis (e.g., 50% or 70%), and recording whether carotid revascularisation was performed and within what time interval.

### Methodological Considerations

Conducting a scoping review and a Delphi consultation process with stakeholders involves several methodological considerations. The identification of national stroke registries and audits was complicated at times by ambiguous and inconsistent reporting on coverage, continuous data collection, and items collected. Many registries, while very comprehensive and with the ambition or potential to be national audits, lacked full national coverage (e.g., ESO RES-Q (91), Indian stroke audit (92), and Bigdata Observatory Platform for Stroke in China (93)), were collected modularly (51), or had shifted from continuous or national to periodic or regional collection due to funding constraints and organisational barriers (53, 54). Data dictionaries and item documentation were often unavailable and had to be compiled from multiple sources, with inconsistent definitions and variable response sets further challenging data charting. To mitigate these issues, data governance for national audits should ensure detailed reporting of registry data quality, item definitions, active status, and coverage to facilitate international benchmarking and performance review. We attempted to reduce selection bias by contacting authors for additional clarifications on audit eligibility and translating data dictionaries to English, where necessary.

In the Delphi consultation, we aimed to reduce potential biases such as dominant perspectives and groupthink by allowing participants ample time and opportunities to vote anonymously offline, review controlled feedback, and change their votes. Adhering to best practices (49, 50), we conducted three rounds of anonymous voting, held Steering Group and Governance Committee meetings to facilitate group discussion, and set *a priori* thresholds for consensus. The Delphi process is inherently subjective, reliant on the representativeness of the expert panel. Our expert group included a diverse range of stakeholders, such as stroke physicians, audit and policy experts, patient representatives, and allied healthcare practitioners, achieving good response rates across voting rounds. However, future research could benefit from a larger and more diverse panel at a national level, but also at European and international levels, to ensure that the individual national audits align with local healthcare and national priorities, as well as adhering to evolving international best practice standards and guidelines.

## Conclusion

This scoping review and Delphi Consensus process identified the “ideal” core outcomes dataset for acute stroke care in Ireland, derived from comprehensive international benchmarking and stakeholder input. The proposed additional items underscore critical gaps, such as the need for more information on risk factor identification, secondary prevention, and the assessment of cognitive outcomes post-stroke. Implementing an ideal dataset with expanded items requires careful consideration of the resource implications, especially for clinical nurse specialists who are often responsible for audit data entry (16). These efforts must also align with best practice outlined by clinical guidelines, emphasising the need for dedicated resources to support audit activities to ensure effective and high-quality data collection that drives continuous quality improvement. Periodic review of audit items is important to ensure data collection aligns with current guidelines and practice at all times, and to facilitate further refinement to enhance item clarity, where necessary. This includes rewording, expanding response options, and organising items with logical sequencing and skip logic. The INAS Governance Committee has begun the process of implementing these recommendations, with close consideration of resourcing implications arising from changes to the audit. Working in partnership with the multi-stakeholder project Steering Group and INAS Governance Committee has ensured that there is a rapid translation of the research findings to audit policy and practice.

In conclusion, the dataset will serve as the “gold standard” for monitoring acute stroke care in Ireland, enhancing patient recovery, and supporting quality improvement initiatives at both local and national levels in Ireland. The present research offers a robust foundation for understanding acute stroke care and audit processes on an international scale. Further evaluation is necessary to assess whether additional audit items enhance its comprehensiveness and contribute to service improvements and, ultimately, enhanced patient outcomes.

## Data Availability

The data that support the findings of this study are available from the corresponding author, [CNM], upon reasonable request.

## Acknowledgements

The authors extend their gratitude to the healthcare professionals, stroke and health services researchers, patient representatives, and personnel working in national stroke policy, patient safety, and service planning who generously contributed their time and invaluable insights to the Delphi consultation. Such collaborative input was instrumental in shaping the outcomes of this study and advancing the project deliverables. We would also like to acknowledge and thank Genevieve Holmes for her support with abstract screening during the review stage.

## Source of Funding

Health Research Board Ireland [APA-2019-036]. The funders had no role in study design, data collection and analysis, decision to publish, or preparation of the manuscript.

### List of tables and figures

- **Table 1**: Final Consensus “Ideal” Core Minimum Dataset for Acute Stroke Care in Ireland
- **Figure 1**. PRISMA flow diagram detailing the scoping review search strategy, the number of identified citations screened, reviewed and excluded, and number of audits included.

### Supplementary Files

- **Supplemental File A:** Electronic Database Search Strategy
- **Supplemental File B:** Detailed Voting Instructions for the Three-Round Delphi Consultation
- **Supplemental File C:** Characteristics of included national and continuous stroke audits and registries.
- **Supplemental File D.** Phase 1 Benchmarking: Comparison of INAS Core Clinical and Thrombectomy Items against International Stroke Audits and Registries
- **Supplemental File E.** Phase 2 Benchmarking: Frequency Counts of International Acute Stroke Care Audit Items, not collected by INAS
- **Supplemental File F.** Results of the three-round Delphi Consultation regarding the retention of the INAS Core Clinical/Thrombectomy Items as part of the “Ideal” Core Minimum Acute Stroke Care Dataset for INAS
- **Supplemental File G.** Results of the three-round Delphi Consultation regarding the recommended inclusion of frequently collected international Items and additional expert-suggested items as part of the “Ideal” Core Minimum Acute Stroke Care Dataset for INAS
- **Supplemental File H.** PRISMA Scoping review checklist

## Footnotes

a Twenty-four INAS Governance Committee and project Steering Group members were invited to take part in the stakeholder consultation. Following Round 1 of the Delphi process, one Governance Committee member left the group (*n*= 1, working in stroke policy); their Round 1 results were tallied and aggregated with the group participant data, but they are excluded from subsequent rounds. An additional project Steering Group member (*n*= 1 researcher) responded to the first two Delphi rounds, and then temporarily left the group and so they were excluded from the final round of the Delphi consultation; their results are counted for Rounds 1 and 2. Hence, in Round 3, 22 eligible stakeholders were consulted.

b There was an additional physiotherapist in the committee, but they hold an academic professorial appointment and hence they are counted as a stroke and health services researcher.

c During the initial search, the Israeli National Stroke Registry may have been conflated with the National Acute Stroke Israeli Survey which is conducted triennially and is therefore non-continuous and was excluded.

