## Supplementary material for "Development of a National Stroke Audit to Enhance the Quality of Acute Stroke Care in Ireland: A Scoping Review and Delphi Consensus Synthesis": Table 1, Figure 1

### Tables & Figures

**Table 1:** Final Consensus "Ideal" Core Minimum Dataset for Acute Stroke Care in Ireland

| | ORIGINAL AUDIT SOURCE | ACUTE STROKE CARE ITEM |
|---|---|---|
| **1** | HIPE | Age |
| **2** | HIPE | Date of discharge |
| **3** | HIPE | Hospital acquired diagnosis |
| **4** | HIPE | Length of stay |
| **5** | HIPE | Principal diagnosis |
| **6** | HIPE | Sex |
| **7** | HIPE | Source of admission – 1 home, 2 transfer from nursing home, convalescent home or other long-stay accommodation, 3 transfer of admitted patient from acute hospital, 4 transfer from non-acute Hospital, 5 transfer from hospice, 6 transfer from psychiatric hospital/unit, 7 new born, 8 temporary place of residence, 9 prison, 0 other |
| **8** | HIPE | Type of admission – 1 elective, 2 elective readmission, 4 emergency, 5 emergency readmission, 6 maternity, 7 new born |
| **9** | HIPE | 1st additional diagnosis |
| **10** | HIPE | Hospital code list |
| **11** | Core Clinical | Which hospital was patient transferred from (if any) |
| **12** | Core Clinical | Why was the patient transferred? – 1 Thrombolysis, 2 Thrombectomy, 3 Neuro Surgery, 8 Other |
| **13** | Core Clinical | Symptom onset date |
| **14** | Core Clinical | Symptom onset time |
| **15** | Core Clinical | If symptom onset date is unknown, what date was the patient last known to be well |
| **16** | Core Clinical | If symptom onset time is unknown, what time was the patient last known to be well |
| **17** | Core Clinical | Did the stroke occur while the patient was in hospital for treatment of another condition – 1 Yes, 2 No, 9 Unknown |
| **18** | Core Clinical | If no, date of presentation to hospital |
| **19** | Core Clinical | If no, time of presentation to hospital |
| **20** | Core Clinical | Brain CT or MRI performed – 1 Yes, 2 No, 3 Performed pre-admission/ hospital transfer, 9 unknown |
| **21** | Core Clinical | If yes, First Brain Imaging date |
| **22** | Core Clinical | If yes, First Brain Imaging time |
| **23** | Core Clinical | Did the patient receive I.V. Thrombolysis – 1 Yes, 2 No, 5 Contraindicated |
| **24** | Core Clinical | If yes, enter date |

| | | |
|---|---|---|
| **25** | Core Clinical | If yes, enter time |
| **26** | Core Clinical | If yes, was intracerebral bleed seen on scan within 36 hrs – 1 Yes, 2 No, 9 Unknown |
| **27** | Core Clinical | If intracerebral bleed, was neuro deterioration associated with it – 1 Yes, 2 No, 9 Unknown |
| **28** | Core Clinical | Did the patient have thrombectomy in this hospital (Beaumont / CUH only) – 1 Yes, 2 No |
| **29** | Core Clinical | Was a swallow screen completed – 1 Yes, 2 No, 9 Unknown |
| **30** | Core Clinical | If yes, was swallow screen completed within 4 hours of presentation – 1 Yes, 2 No, 9 Unknown |
| **31** | Core Clinical | Modified Rankin Scale pre-stroke – 0 Zero, 1 One, 2 Two, 3 Three, 4 Four, 5 Five, 6 Six, 9 Unknown |
| **32** | Core Clinical | Admitted to Stroke Unit (Key Performance Indicator) – 1 Yes, 2 No |
| **33** | Core Clinical | If yes, date admitted to Stroke Unit (Key Performance Indicator) |
| **34** | Core Clinical | If yes, date discharged from Stroke Unit (Key Performance Indicator) |
| **35** | Core Clinical | If no, reason why – 1 No stroke unit, 2 Bed not available, 5 Infection control risk, 8 Other |
| **36** | Core Clinical | Allied Health Professional (AHP) Assessment – 1 Yes, 2 No |
| **37** | Core Clinical | If yes, Physiotherapy – 1 Yes, 2 No |
| **38** | Core Clinical | If yes, Occupational Therapy – 1 Yes, 2 No |
| **39** | Core Clinical | If yes, Speech and Language Therapy – 1 Yes, 2 No |
| **40** | Core Clinical | If yes, Dietetics – 1 Yes, 2 No |
| **41** | Core Clinical | If yes, Medical Social Worker – 1 Yes, 2 No |
| **42** | Core Clinical | If yes, Psychology – 1 Yes, 2 No |
| **43** | Core Clinical | Was the patient assessed by Stroke Nurse Specialist – 1 Yes, 2 No, 9 Unknown |
| **44** | Core Clinical | Was an assessment of mood completed and documented by a member of the multidisciplinary team – 1 Yes, 2 No, 3 Not indicated, 9 Unknown |
| **45** | Core Clinical | New or Altered Antithrombotic Therapy prescribed for acute treatment – 1 Yes, 2 No, 3 Contraindicated, 9 Unknown |
| **46** | Core Clinical | If yes, Antiplatelet or Anticoagulant (for acute treatment) start date |
| **47** | Core Clinical | Does the patient have Atrial Fibrillation – 1 Yes, 2 No, 4 Results pending, 9 Unknown |
| **48** | Core Clinical | If Atrial Fibrillation, was atrial fibrillation known prior to stroke onset – 1 Yes, 2 No, 9 Unknown |
| **49** | Core Clinical | If atrial fibrillation known prior to stroke onset, was Antiplatelet and/or Anticoagulant prescribed prior to Stroke onset – 1 Yes, 2 No, 9 Unknown |
| **50** | Core Clinical | If yes, please specify Antiplatelet / Anticoagulant - prior to stroke - 0 NOAC, 1 Warfarin, 5 Aspirin, 6 Clopidogrel, 7 Other Antiplatelet, 8 Dual Antiplatelet Therapy, 9 Antiplatelet & Anticoagulant |
| **51** | Core Clinical | If atrial fibrillation known prior to stroke onset, and on Warfarin, was INR (International Normalised Ratio) 2-3 at Stroke onset – 1 Yes, 2 No, 9 Unknown |
| **52** | Core Clinical | If Atrial Fibrillation, Anticoagulation prescribed for secondary prevention – 1 Yes, 2 No, 9 Unknown |
| **53** | Core Clinical | If yes, please specify Antiplatelet / Anticoagulant - on discharge - 0 NOAC, 1 Warfarin, 5 Aspirin, 6 Clopidogrel, 7 Other Antiplatelet, 8 Dual Antiplatelet Therapy, 9 Antiplatelet & Anticoagulant |
| **54** | Core Clinical | If no, please enter reason documented - 1 No reason documented, 2 Major bleeding (prior history), 3 Severe illness (e.g. cancer, dementia), 4 Poor compliance (known or suspected), 5 Patient refused anticoagulation, 6 Alcohol excess, 7 Falls, 8 Extreme frailty, 9 Liver disease |
| **55** | Core Clinical | Modified Rankin Scale on discharge - 0 Zero, 1 One, 2 Two, 3 Three, 4 Four, 5 Five, 6 Six, 9 Unknown |
| **56** | Core Clinical | Discharge destination - 1 Home, 2 Patient died, 3 Discharge to long term care, 4 Discharge to off-site rehab, 5 Transfer to referring hospital, 6 Transfer to other hospital for on-going stroke care, 7 Home with ESD, 8 Other, 9 Unknown |
| **57** | Thrombectomy | NIHSS pre-thrombectomy |
| **58** | Thrombectomy | Date of performance of non-contrast CT |
| **59** | Thrombectomy | Time of performance of non-contrast CT |
| **60** | Thrombectomy | Date of performance of non-contrast CTA |

| | | |
|---|---|---|
| **61** | Thrombectomy | Time of performance of non-contrast CTA |
| **62** | Thrombectomy | Date of contact with the endovascular stroke centre |
| **63** | Thrombectomy | Time of contact with the endovascular stroke centre |
| **64** | Thrombectomy | Date of decision to transfer patient |
| **65** | Thrombectomy | Time of decision to transfer patient |
| **66** | Thrombectomy | Was the patient transferred from another hospital - 1 Yes, 2 No, 9 Unknown |
| **67** | Thrombectomy | If yes, what date did the patient arrive at the referring/first hospital |
| **68** | Thrombectomy | If yes, what time did the patient arrive at the referring/first hospital |
| **69** | Thrombectomy | If yes, what date did the patient leave the referring/first hospital for transfer to the EVT centre? |
| **70** | Thrombectomy | If yes, what time did the patient leave the referring/first hospital for transfer to the EVT centre? |
| **71** | Thrombectomy | Date of arrival at the endovascular stroke centre |
| **72** | Thrombectomy | Time of arrival at the endovascular stroke centre |
| **73** | Thrombectomy | Did the patient have repeat non-invasive imaging in the endovascular stroke centre - 1 Yes, 2 No, 9 Unknown |
| **74** | Thrombectomy | If yes, please specify - 1 Non-contrast CT, 2 CTA, 3 Perfusion CT, 4 MRI |
| **75** | Thrombectomy | Site of most proximal occlusion - 1 MCA 1, 2 MCA 2, 3 Basilar, 4 ICA carotid T/L, 5 ICA cervical segment, 6 PCA, 7 Vertebro basilar |
| **76** | Thrombectomy | Second occlusion site |
| **77** | Thrombectomy | Associated carotid stenosis greater than 50% - 1 Yes, 2 No, 9 Unknown |
| **78** | Thrombectomy | TICI pre thrombectomy |
| **79** | Thrombectomy | TICI post thrombectomy |
| **80** | Thrombectomy | Date of groin puncture |
| **81** | Thrombectomy | Time of groin puncture |
| **82** | Thrombectomy | Date of first pass |
| **83** | Thrombectomy | Time of first pass |
| **84** | Thrombectomy | Date of first reperfusion |
| **85** | Thrombectomy | Time of first reperfusion |
| **86** | Thrombectomy | Date of final angio |
| **87** | Thrombectomy | Time of final angio |
| **88** | Thrombectomy | Immediate complications - Not Applicable, 1 Haemorrhage, 2 Embolus into separate vascular territory, 3 Dissection, 8 Other, 9 Unknown |
| **89** | Thrombectomy | NIHSS 24-hour post-thrombectomy |
| **90** | Thrombectomy | Following procedure was patient transferred immediately back to primary receiving hospital - 1 Yes, 2 No, 9 Unknown |
| **91** | Thrombectomy | If no, when was patient admitted to the endovascular stroke centre- 1 0-3 hrs, 2 3-12 hrs, 3 12-24 hrs, 4 24+ hrs |
| **92** | International | Patient Record ID [Patient Details] |
| **93** | International | Did the patient arrive by ambulance? [Admission Details] |
| **94** | International | Previous stroke (A history of stroke prior to this current episode, excluding TIAs) [History of Known Risk Factors] |
| **95** | International | Previous TIA (Transient Ischemic Attack (TIA) or mini stroke) [History of Known Risk Factors] |
| **96** | International | Diabetes [History of Known Risk Factors] |
| **97** | International | Hypercholesterolaemia [History of Known Risk Factors] |
| **98** | International | Hypertension [History of Known Risk Factors] |
| **99** | International | Recent myocardial infarction [History of Known Risk Factors] |
| **100** | International | Ischaemic heart disease [History of Known Risk Factors] |
| **101** | International | High alcohol consumption [History of Known Risk Factors] |

| | | |
|---|---|---|
| **102** | International | Current smoker [History of Known Risk Factors] |
| **103** | International | Past smoker [History of Known Risk Factors] |
| **104** | International | Obesity [History of Known Risk Factors] |
| **105** | International | Peripheral vascular disease [History of Known Risk Factors] |
| **106** | International | Did the patient take antihypertensives prior to this stroke? [Pre-Admission Medication] |
| **107** | International | Did the patient take lipid lowering treatment prior to this stroke? [Pre-Admission Medication] |
| **108** | International | NIHSS at baseline [Acute Clinical Data] |
| **109** | International | Aspiration pneumonia [Complications during Hospital Admission] |
| **110** | International | Deep Vein Thrombosis (DVT) [Complications during Hospital Admission] |
| **111** | International | Pulmonary Thrombo-Embolism (PE) [Complications during Hospital Admission] |
| **112** | International | New stroke [Complications during Hospital Admission] |
| **113** | International | Urinary tract infection [Complications during Hospital Admission] |
| **114** | International | Seizures [Complications during Hospital Admission] |
| **115** | International | On discharge was the patient prescribed antihypertensive agents? [Medication Prescribed at Discharge] |
| **116** | Additional expert-suggested | Location of Intracerebral haematoma in ICH. |
| **117** | Additional expert-suggested | mRS at 90 days |
| **118** | Additional expert-suggested | Was screening for cognitive impairment completed using a valid screening measure? 1 Yes; 2 No; 3 Unable to complete due to patient factors; 9 Unknown |
| **119** | Additional expert-suggested | Was screening for aphasia conducted? |
| **120** | Additional expert-suggested | Does the patient have aphasia? |
| **121** | Additional expert-suggested | Dementia (diagnosis of pre-stroke dementia) |

***Note.*** Abbreviations: Angio: angiogram; CT: computed tomography; CTA: computed tomography angiography; CUH: Cork University Hospital; ESD: early supported discharge; EVT: endovascular thrombectomy; HIPE: Hospital In-Patient Enquiry; ICA: internal carotid artery; IV: intravenous; MCA: middle cerebral artery; MRI: magnetic resonance imaging; mRS: modified Rankin Scake; NIHSS: National Institutes of Health Stroke Scale; NOAC: non-vitamin K oral anticoagulants; PCA: posterior cerebral artery; TIA: transient ischaemic attack; TICI: thrombolysis in cerebral infarction.

**Figure 1.** PRISMA flow diagram detailing the scoping review search strategy, the number of identified citations screened, reviewed and excluded, and number of audits included.

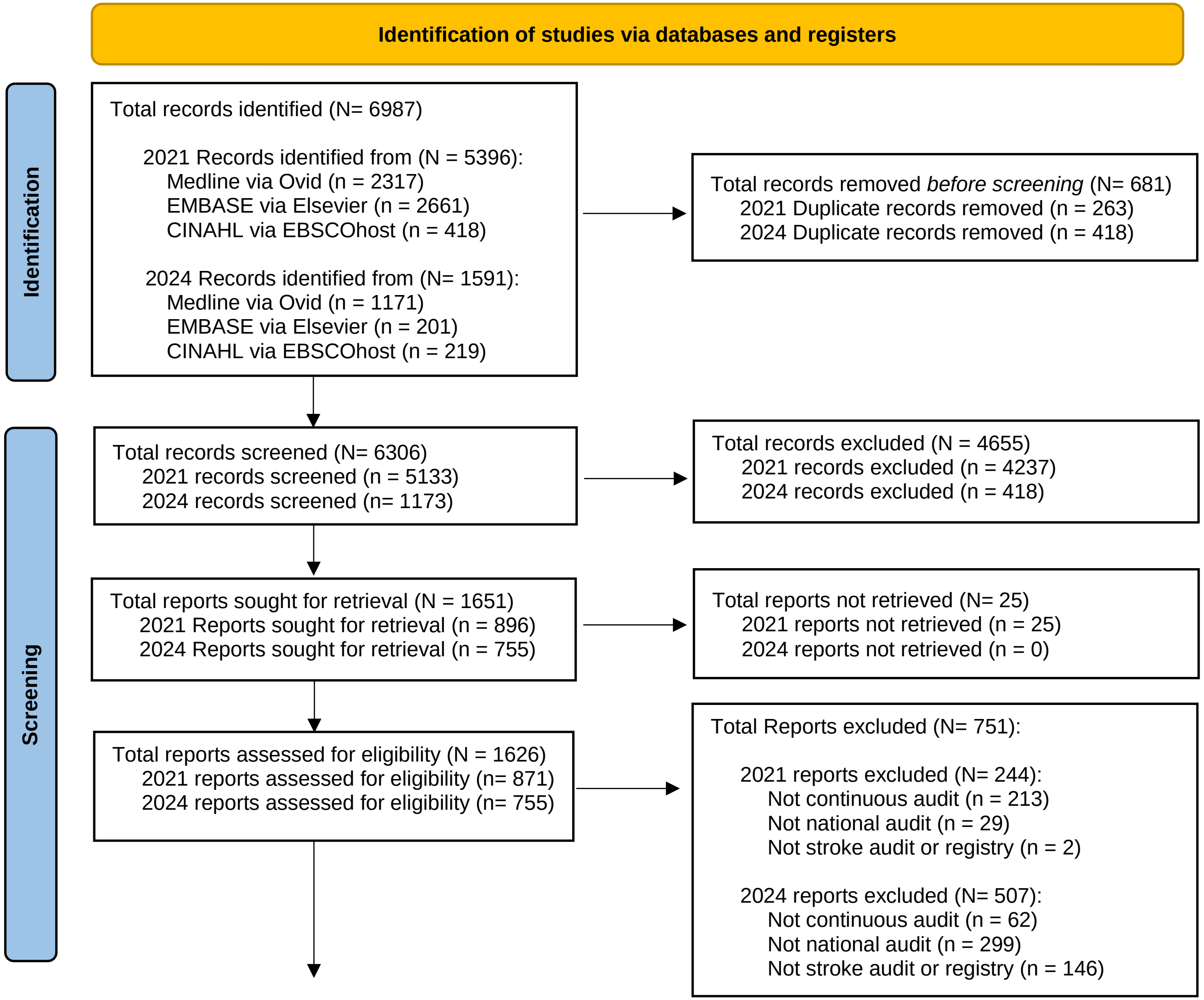

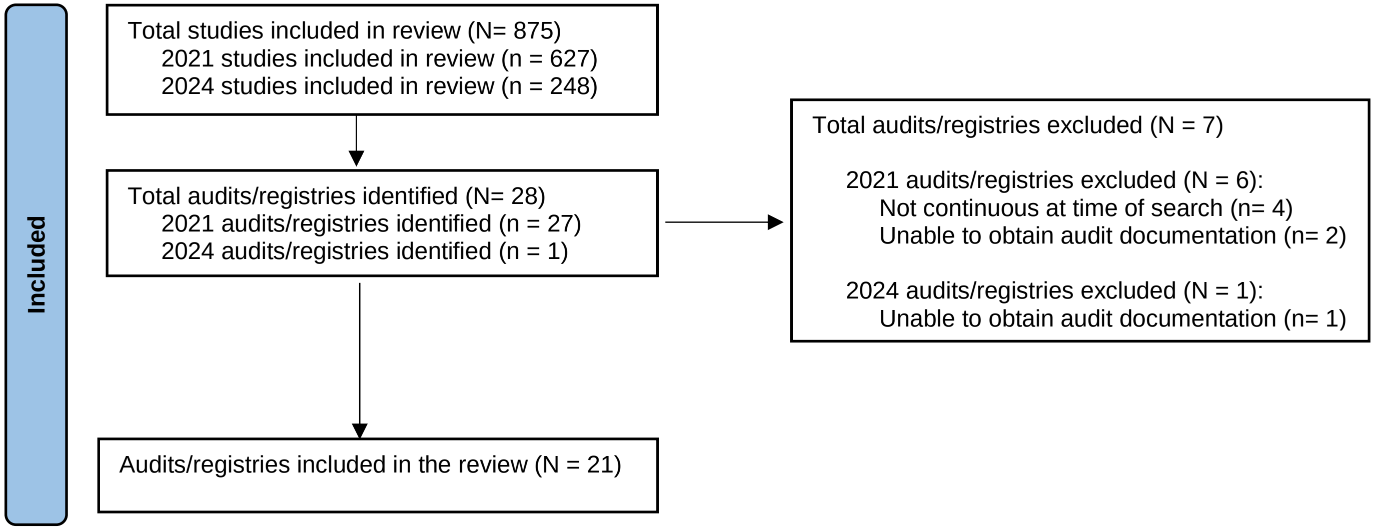
Included
Total studies included in review (N= 875)
2021 studies included in review (n = 627)
2024 studies included in review (n = 248)
Total audits/registries identified (N= 28)
2021 audits/registries identified (n = 27)
2024 audits/registries identified (n = 1)
Total audits/registries excluded (N = 7)
2021 audits/registries excluded (N = 6):
Not continuous at time of search (n= 4)
Unable to obtain audit documentation (n= 2)
2024 audits/registries excluded (N = 1):
Unable to obtain audit documentation (n= 1)
Audits/registries included in the review (N = 21)
