## supplemental files for "Development of a National Stroke Audit to Enhance the Quality of Acute Stroke Care in Ireland: A Scoping Review and Delphi Consensus Synthesis"

### Tables & Figures

**Table 1:** Final Consensus "Ideal" Core Minimum Dataset for Acute Stroke Care in Ireland

| | ORIGINAL AUDIT SOURCE | ACUTE STROKE CARE ITEM |
|---|---|---|
| **1** | HIPE | Age |
| **2** | HIPE | Date of discharge |
| **3** | HIPE | Hospital acquired diagnosis |
| **4** | HIPE | Length of stay |
| **5** | HIPE | Principal diagnosis |
| **6** | HIPE | Sex |
| **7** | HIPE | Source of admission – 1 home, 2 transfer from nursing home, convalescent home or other long-stay accommodation, 3 transfer of admitted patient from acute hospital, 4 transfer from non-acute Hospital, 5 transfer from hospice, 6 transfer from psychiatric hospital/unit, 7 new born, 8 temporary place of residence, 9 prison, 0 other |
| **8** | HIPE | Type of admission – 1 elective, 2 elective readmission, 4 emergency, 5 emergency readmission, 6 maternity, 7 new born |
| **9** | HIPE | 1st additional diagnosis |
| **10** | HIPE | Hospital code list |
| **11** | Core Clinical | Which hospital was patient transferred from (if any) |
| **12** | Core Clinical | Why was the patient transferred? – 1 Thrombolysis, 2 Thrombectomy, 3 Neuro Surgery, 8 Other |
| **13** | Core Clinical | Symptom onset date |
| **14** | Core Clinical | Symptom onset time |
| **15** | Core Clinical | If symptom onset date is unknown, what date was the patient last known to be well |
| **16** | Core Clinical | If symptom onset time is unknown, what time was the patient last known to be well |
| **17** | Core Clinical | Did the stroke occur while the patient was in hospital for treatment of another condition – 1 Yes, 2 No, 9 Unknown |
| **18** | Core Clinical | If no, date of presentation to hospital |
| **19** | Core Clinical | If no, time of presentation to hospital |
| **20** | Core Clinical | Brain CT or MRI performed – 1 Yes, 2 No, 3 Performed pre-admission/ hospital transfer, 9 unknown |
| **21** | Core Clinical | If yes, First Brain Imaging date |
| **22** | Core Clinical | If yes, First Brain Imaging time |
| **23** | Core Clinical | Did the patient receive I.V. Thrombolysis – 1 Yes, 2 No, 5 Contraindicated |
| **24** | Core Clinical | If yes, enter date |

| | | |
|---|---|---|
| **25** | Core Clinical | If yes, enter time |
| **26** | Core Clinical | If yes, was intracerebral bleed seen on scan within 36 hrs – 1 Yes, 2 No, 9 Unknown |
| **27** | Core Clinical | If intracerebral bleed, was neuro deterioration associated with it – 1 Yes, 2 No, 9 Unknown |
| **28** | Core Clinical | Did the patient have thrombectomy in this hospital (Beaumont / CUH only) – 1 Yes, 2 No |
| **29** | Core Clinical | Was a swallow screen completed – 1 Yes, 2 No, 9 Unknown |
| **30** | Core Clinical | If yes, was swallow screen completed within 4 hours of presentation – 1 Yes, 2 No, 9 Unknown |
| **31** | Core Clinical | Modified Rankin Scale pre-stroke – 0 Zero, 1 One, 2 Two, 3 Three, 4 Four, 5 Five, 6 Six, 9 Unknown |
| **32** | Core Clinical | Admitted to Stroke Unit (Key Performance Indicator) – 1 Yes, 2 No |
| **33** | Core Clinical | If yes, date admitted to Stroke Unit (Key Performance Indicator) |
| **34** | Core Clinical | If yes, date discharged from Stroke Unit (Key Performance Indicator) |
| **35** | Core Clinical | If no, reason why – 1 No stroke unit, 2 Bed not available, 5 Infection control risk, 8 Other |
| **36** | Core Clinical | Allied Health Professional (AHP) Assessment – 1 Yes, 2 No |
| **37** | Core Clinical | If yes, Physiotherapy – 1 Yes, 2 No |
| **38** | Core Clinical | If yes, Occupational Therapy – 1 Yes, 2 No |
| **39** | Core Clinical | If yes, Speech and Language Therapy – 1 Yes, 2 No |
| **40** | Core Clinical | If yes, Dietetics – 1 Yes, 2 No |
| **41** | Core Clinical | If yes, Medical Social Worker – 1 Yes, 2 No |
| **42** | Core Clinical | If yes, Psychology – 1 Yes, 2 No |
| **43** | Core Clinical | Was the patient assessed by Stroke Nurse Specialist – 1 Yes, 2 No, 9 Unknown |
| **44** | Core Clinical | Was an assessment of mood completed and documented by a member of the multidisciplinary team – 1 Yes, 2 No, 3 Not indicated, 9 Unknown |
| **45** | Core Clinical | New or Altered Antithrombotic Therapy prescribed for acute treatment – 1 Yes, 2 No, 3 Contraindicated, 9 Unknown |
| **46** | Core Clinical | If yes, Antiplatelet or Anticoagulant (for acute treatment) start date |
| **47** | Core Clinical | Does the patient have Atrial Fibrillation – 1 Yes, 2 No, 4 Results pending, 9 Unknown |
| **48** | Core Clinical | If Atrial Fibrillation, was atrial fibrillation known prior to stroke onset – 1 Yes, 2 No, 9 Unknown |
| **49** | Core Clinical | If atrial fibrillation known prior to stroke onset, was Antiplatelet and/or Anticoagulant prescribed prior to Stroke onset – 1 Yes, 2 No, 9 Unknown |
| **50** | Core Clinical | If yes, please specify Antiplatelet / Anticoagulant - prior to stroke - 0 NOAC, 1 Warfarin, 5 Aspirin, 6 Clopidogrel, 7 Other Antiplatelet, 8 Dual Antiplatelet Therapy, 9 Antiplatelet & Anticoagulant |
| **51** | Core Clinical | If atrial fibrillation known prior to stroke onset, and on Warfarin, was INR (International Normalised Ratio) 2-3 at Stroke onset – 1 Yes, 2 No, 9 Unknown |
| **52** | Core Clinical | If Atrial Fibrillation, Anticoagulation prescribed for secondary prevention – 1 Yes, 2 No, 9 Unknown |
| **53** | Core Clinical | If yes, please specify Antiplatelet / Anticoagulant - on discharge - 0 NOAC, 1 Warfarin, 5 Aspirin, 6 Clopidogrel, 7 Other Antiplatelet, 8 Dual Antiplatelet Therapy, 9 Antiplatelet & Anticoagulant |
| **54** | Core Clinical | If no, please enter reason documented - 1 No reason documented, 2 Major bleeding (prior history), 3 Severe illness (e.g. cancer, dementia), 4 Poor compliance (known or suspected), 5 Patient refused anticoagulation, 6 Alcohol excess, 7 Falls, 8 Extreme frailty, 9 Liver disease |
| **55** | Core Clinical | Modified Rankin Scale on discharge - 0 Zero, 1 One, 2 Two, 3 Three, 4 Four, 5 Five, 6 Six, 9 Unknown |
| **56** | Core Clinical | Discharge destination - 1 Home, 2 Patient died, 3 Discharge to long term care, 4 Discharge to off-site rehab, 5 Transfer to referring hospital, 6 Transfer to other hospital for on-going stroke care, 7 Home with ESD, 8 Other, 9 Unknown |
| **57** | Thrombectomy | NIHSS pre-thrombectomy |
| **58** | Thrombectomy | Date of performance of non-contrast CT |
| **59** | Thrombectomy | Time of performance of non-contrast CT |
| **60** | Thrombectomy | Date of performance of non-contrast CTA |

| | | |
|---|---|---|
| **61** | Thrombectomy | Time of performance of non-contrast CTA |
| **62** | Thrombectomy | Date of contact with the endovascular stroke centre |
| **63** | Thrombectomy | Time of contact with the endovascular stroke centre |
| **64** | Thrombectomy | Date of decision to transfer patient |
| **65** | Thrombectomy | Time of decision to transfer patient |
| **66** | Thrombectomy | Was the patient transferred from another hospital - 1 Yes, 2 No, 9 Unknown |
| **67** | Thrombectomy | If yes, what date did the patient arrive at the referring/first hospital |
| **68** | Thrombectomy | If yes, what time did the patient arrive at the referring/first hospital |
| **69** | Thrombectomy | If yes, what date did the patient leave the referring/first hospital for transfer to the EVT centre? |
| **70** | Thrombectomy | If yes, what time did the patient leave the referring/first hospital for transfer to the EVT centre? |
| **71** | Thrombectomy | Date of arrival at the endovascular stroke centre |
| **72** | Thrombectomy | Time of arrival at the endovascular stroke centre |
| **73** | Thrombectomy | Did the patient have repeat non-invasive imaging in the endovascular stroke centre - 1 Yes, 2 No, 9 Unknown |
| **74** | Thrombectomy | If yes, please specify - 1 Non-contrast CT, 2 CTA, 3 Perfusion CT, 4 MRI |
| **75** | Thrombectomy | Site of most proximal occlusion - 1 MCA 1, 2 MCA 2, 3 Basilar, 4 ICA carotid T/L, 5 ICA cervical segment, 6 PCA, 7 Vertebro basilar |
| **76** | Thrombectomy | Second occlusion site |
| **77** | Thrombectomy | Associated carotid stenosis greater than 50% - 1 Yes, 2 No, 9 Unknown |
| **78** | Thrombectomy | TICI pre thrombectomy |
| **79** | Thrombectomy | TICI post thrombectomy |
| **80** | Thrombectomy | Date of groin puncture |
| **81** | Thrombectomy | Time of groin puncture |
| **82** | Thrombectomy | Date of first pass |
| **83** | Thrombectomy | Time of first pass |
| **84** | Thrombectomy | Date of first reperfusion |
| **85** | Thrombectomy | Time of first reperfusion |
| **86** | Thrombectomy | Date of final angio |
| **87** | Thrombectomy | Time of final angio |
| **88** | Thrombectomy | Immediate complications - Not Applicable, 1 Haemorrhage, 2 Embolus into separate vascular territory, 3 Dissection, 8 Other, 9 Unknown |
| **89** | Thrombectomy | NIHSS 24-hour post-thrombectomy |
| **90** | Thrombectomy | Following procedure was patient transferred immediately back to primary receiving hospital - 1 Yes, 2 No, 9 Unknown |
| **91** | Thrombectomy | If no, when was patient admitted to the endovascular stroke centre- 1 0-3 hrs, 2 3-12 hrs, 3 12-24 hrs, 4 24+ hrs |
| **92** | International | Patient Record ID [Patient Details] |
| **93** | International | Did the patient arrive by ambulance? [Admission Details] |
| **94** | International | Previous stroke (A history of stroke prior to this current episode, excluding TIAs) [History of Known Risk Factors] |
| **95** | International | Previous TIA (Transient Ischemic Attack (TIA) or mini stroke) [History of Known Risk Factors] |
| **96** | International | Diabetes [History of Known Risk Factors] |
| **97** | International | Hypercholesterolaemia [History of Known Risk Factors] |
| **98** | International | Hypertension [History of Known Risk Factors] |
| **99** | International | Recent myocardial infarction [History of Known Risk Factors] |
| **100** | International | Ischaemic heart disease [History of Known Risk Factors] |
| **101** | International | High alcohol consumption [History of Known Risk Factors] |

| | | |
|---|---|---|
| **102** | International | Current smoker [History of Known Risk Factors] |
| **103** | International | Past smoker [History of Known Risk Factors] |
| **104** | International | Obesity [History of Known Risk Factors] |
| **105** | International | Peripheral vascular disease [History of Known Risk Factors] |
| **106** | International | Did the patient take antihypertensives prior to this stroke? [Pre-Admission Medication] |
| **107** | International | Did the patient take lipid lowering treatment prior to this stroke? [Pre-Admission Medication] |
| **108** | International | NIHSS at baseline [Acute Clinical Data] |
| **109** | International | Aspiration pneumonia [Complications during Hospital Admission] |
| **110** | International | Deep Vein Thrombosis (DVT) [Complications during Hospital Admission] |
| **111** | International | Pulmonary Thrombo-Embolism (PE) [Complications during Hospital Admission] |
| **112** | International | New stroke [Complications during Hospital Admission] |
| **113** | International | Urinary tract infection [Complications during Hospital Admission] |
| **114** | International | Seizures [Complications during Hospital Admission] |
| **115** | International | On discharge was the patient prescribed antihypertensive agents? [Medication Prescribed at Discharge] |
| **116** | Additional expert-suggested | Location of Intracerebral haematoma in ICH. |
| **117** | Additional expert-suggested | mRS at 90 days |
| **118** | Additional expert-suggested | Was screening for cognitive impairment completed using a valid screening measure? 1 Yes; 2 No; 3 Unable to complete due to patient factors; 9 Unknown |
| **119** | Additional expert-suggested | Was screening for aphasia conducted? |
| **120** | Additional expert-suggested | Does the patient have aphasia? |
| **121** | Additional expert-suggested | Dementia (diagnosis of pre-stroke dementia) |

***Note.*** Abbreviations: Angio: angiogram; CT: computed tomography; CTA: computed tomography angiography; CUH: Cork University Hospital; ESD: early supported discharge; EVT: endovascular thrombectomy; HIPE: Hospital In-Patient Enquiry; ICA: internal carotid artery; IV: intravenous; MCA: middle cerebral artery; MRI: magnetic resonance imaging; mRS: modified Rankin Scake; NIHSS: National Institutes of Health Stroke Scale; NOAC: non-vitamin K oral anticoagulants; PCA: posterior cerebral artery; TIA: transient ischaemic attack; TICI: thrombolysis in cerebral infarction.

**Figure 1.** PRISMA flow diagram detailing the scoping review search strategy, the number of identified citations screened, reviewed and excluded, and number of audits included.

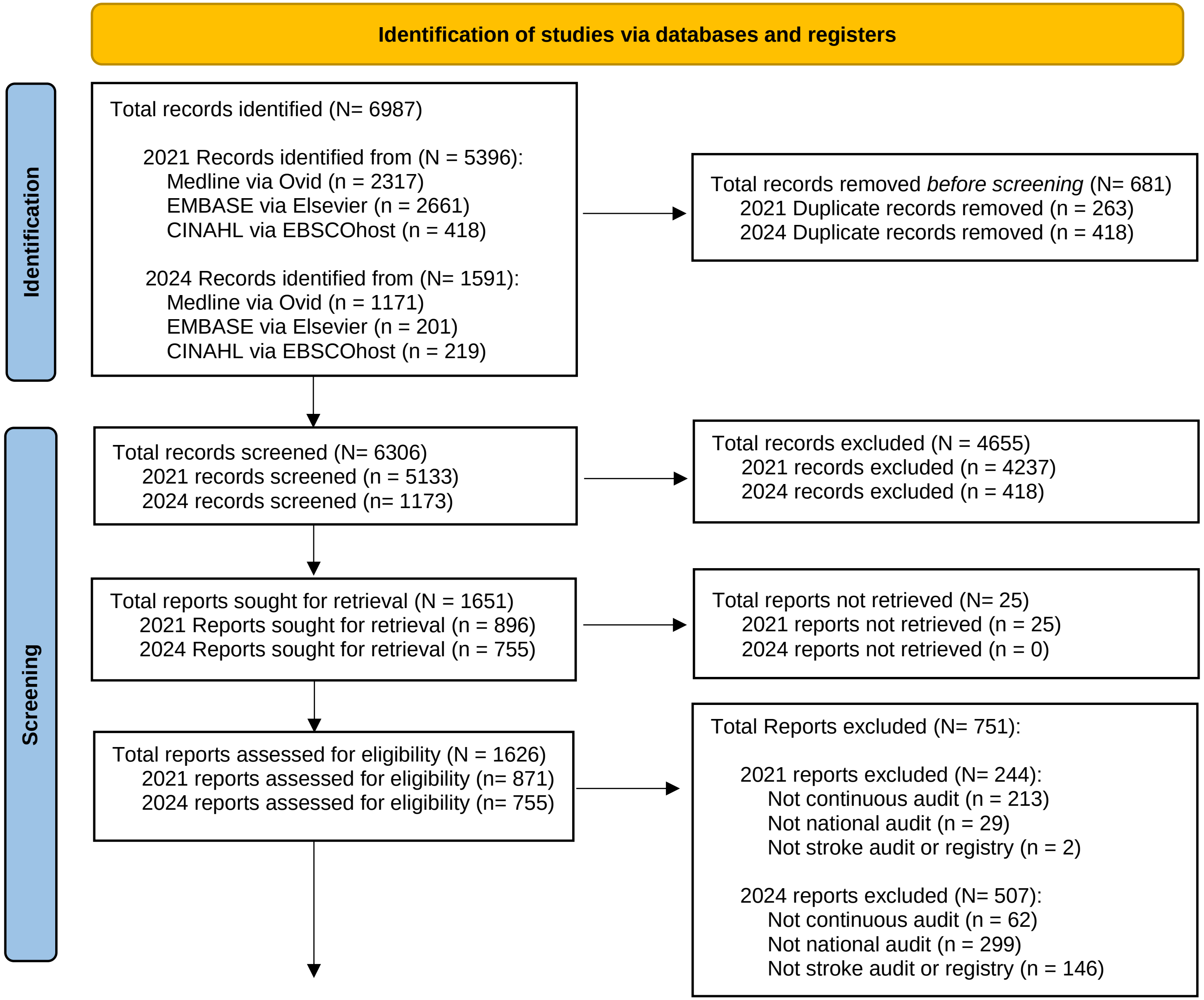

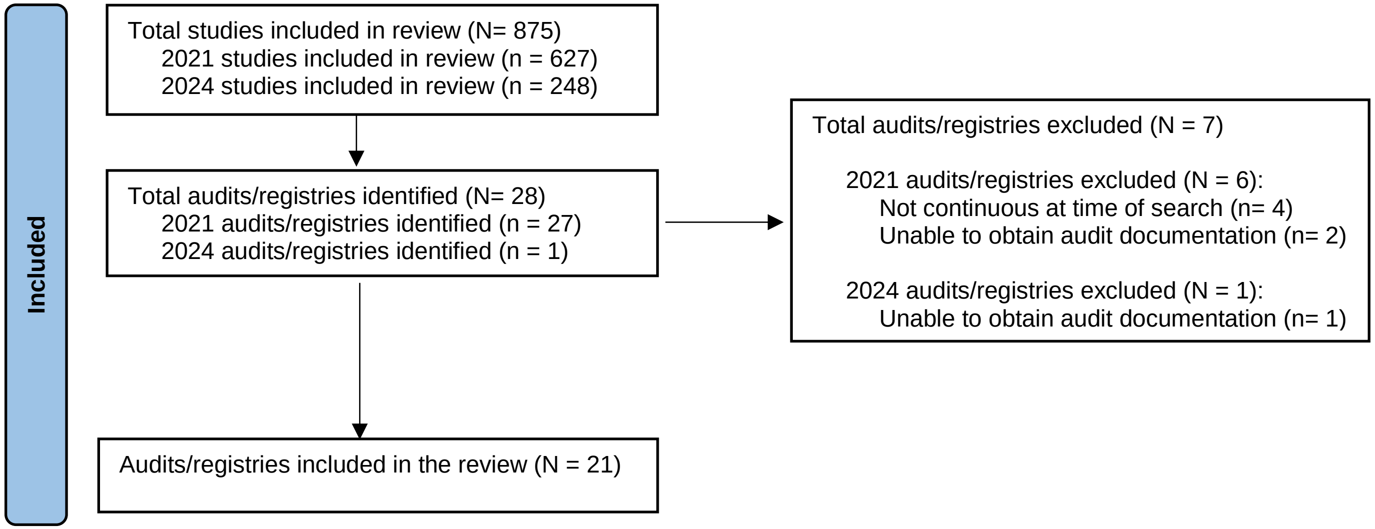
Included
Total studies included in review (N= 875)
2021 studies included in review (n = 627)
2024 studies included in review (n = 248)
Total audits/registries identified (N= 28)
2021 audits/registries identified (n = 27)
2024 audits/registries identified (n = 1)
Total audits/registries excluded (N = 7)
2021 audits/registries excluded (N = 6):
Not continuous at time of search (n= 4)
Unable to obtain audit documentation (n= 2)
2024 audits/registries excluded (N = 1):
Unable to obtain audit documentation (n= 1)
Audits/registries included in the review (N = 21)

### Supplemental Files

**Supplemental File A:** Electronic Database Search Strategy

| | EMBASE on Elsevier.com | 2021 Search | 2024 Search |
|---|---|---|---|
| 1 | Stroke:ti,ab OR ‘cerebral hemorrhage’:ti,ab OR 'cerebral haemorrhage':ab,ti OR 'cerebrovascular accident'/exp OR 'brain ischemia'/exp OR ‘brain ischemia’:ti,ab OR ‘brain ischaemia’:ti,ab OR ‘ischemic attack’:ti,ab OR ‘ischaemic attack’:ti,ab | 589960 | 730338 |
| 2 | Stroke NEXT/3 registry OR stroke NEXT/3 registries OR ‘National Stroke Registry’:ti,ab OR ‘National Stroke Register’:ti,ab OR stroke NEXT/3 audit$ | 5133 | 6818 |
| 3 | ((‘quality register’:ti,ab OR ‘quality registry’:ti,ab OR ‘performance indicator*’:ti,ab OR ‘quality indicator*’:ti,ab) AND stroke:ti,ab,de,kw) | 660 | 863 |
| 4 | (Audit NEXT/2 guideline$) OR (registry NEXT/2 guideline$) | 222 | 273 |
| 5 | 2 OR 3 OR 4 | 5906 | 7820 |
| 6 | 1 AND 5 | 5655 | 7515 |
| 7 | #6 AND [2010-2021]/py | 4838 | 5369 |
| 8 | #7 AND [humans]/lim AND [english]/lim | 4393 | 4920 |
| 9 | #8 AND [medline]/lim | 1732 | 1895 |
| 9 | (stroke:ti,ab OR 'cerebral hemorrhage':ti,ab OR 'cerebral haemorrhage':ab,ti OR 'cerebrovascular accident'/exp OR 'brain ischemia'/exp OR 'brain ischemia':ti,ab OR 'brain ischaemia':ti,ab OR 'ischemic attack':ti,ab OR 'ischaemic attack':ti,ab) AND ((stroke NEXT/3 registry) OR (stroke NEXT/3 registries) OR 'national stroke registry':ti,ab OR 'national stroke register':ti,ab OR (stroke NEXT/3 audit$) OR (('quality register':ti,ab OR 'quality registry':ti,ab OR 'performance | N/A | 1895 |

| | | | |
|---|---|---|---|
| | indicator*':ti,ab OR 'quality indicator*':ti,ab) AND stroke:ti,ab,de,kw) OR (audit NEXT/2 guideline$) OR (registry NEXT/2 guideline$)) AND [2010-2021]/py AND [humans]/lim AND [english]/lim AND [medline]/lim | | |
| 10 | #8 NOT #9 | **2661** | N/A |
| 10 | #9 AND [2021-2024]/py | N/A | 226 |
| 11 | #10 AND [01-02-2021]/sd | N/A | 201 |
| 11 | (stroke:ti,ab OR 'cerebral hemorrhage':ti,ab OR 'cerebral haemorrhage':ab,ti OR 'cerebrovascular accident'/exp OR 'brain ischemia'/exp OR 'brain ischemia':ti,ab OR 'brain ischaemia':ti,ab OR 'ischemic attack':ti,ab OR 'ischaemic attack':ti,ab) AND ((stroke NEXT/3 registry) OR (stroke NEXT/3 registries) OR 'national stroke registry':ti,ab OR 'national stroke register':ti,ab OR (stroke NEXT/3 audit$) OR (('quality register':ti,ab OR 'quality registry':ti,ab OR 'performance indicator*':ti,ab OR 'quality indicator*':ti,ab) AND stroke:ti,ab,de,kw) OR (audit NEXT/2 guideline$) OR (registry NEXT/2 guideline$)) AND [2010-2021]/py AND [humans]/lim AND [english]/lim AND [2021-2024]/py AND [medline]/lim AND [01-02-2021]/sd | N/A | **201** |

| | **Ovid MEDLINE(R) and Epub Ahead of Print, In-Process & Other Non-Indexed Citations, Daily and Versions(R) 1946 to February 03, 2021** | **2021 Search** | **2024 Search** |
|---|---|---|---|
| 1 | exp stroke/ OR stroke.mp. OR exp cerebral hemorrhage/ OR cerebral h*morrhage.mp. OR exp brain ischemia/ OR brain isch*mia.mp. OR exp Ischemic Attack, Transient/ OR isch*mic attack.mp. | 398644 | 485503 |
| 2 | ((Stroke adj3 registry) OR (stroke adj3 registries) OR (National Stroke Registry) OR (National Stroke Register) OR (stroke adj3 audit$)).mp. | 2455 | 3372 |
| 3 | (quality register.mp. OR quality registry.mp. OR performance indicator*.mp. OR quality indicator*.mp.) AND stroke.mp. | 698 | 873 |
| 4 | ((Audit adj2 guideline$) OR (registry adj2 guideline$)).mp. | 404 | 505 |
| 5 | 2 OR 3 OR 4 | 3437 | 4591 |
| 6 | 1 AND 5 | 3096 | 4176 |
| 7 | limit 6 to yr="2010 - 2021" | **2317** | N/A |
| 7 | limit 6 to yr="2010 -Current" | N/A | 3397 |
| 8 | limit 7 to yr="2021 -Current" | N/A | **1171** |

| | CINAHL | 2021 Search | 2024 Search |
|---|---|---|---|
| 1 | (MH “Stroke+”) OR (MH “Cerebral Hemorrhage+”) OR (MH “Cerebral Ischemia+”) OR (MH “Cerebral Ischemia, Transient) OR TI stroke OR AB stroke OR TI ‘cerebral haemorrhage’ OR AB ‘cerebral haemorrhage’ OR TI ‘brain ischemia’ OR AB ‘brain ischaemia’ OR TI ‘ischemic attack’ OR AB ‘ischaemic attack’ | 126938 | 147473 |
| 2 | (Stroke N3 registry) OR (stroke N3 registries) OR TX ‘National Stroke Registry’ OR TX ‘National Stroke Register’ OR (stroke N3 audit$) | 1490 | 1792 |
| 3 | (TX ‘quality register’ OR TX ‘quality registry’ OR TX ‘performance indicator$’ OR TX ‘quality indicator$’) AND ( TI stroke OR AB stroke) | 513 | 613 |
| 4 | (Audit N2 guideline$) OR (registry N2 guideline$) | 412 | 492 |
| 5 | 2 or 3 or 4 | 2295 | 2770 |
| 6 | 1 and 5 | 1850 | 2236 |
| 7 | Limit 6 to 2010 to 2021 | 1489 | N/A |
| 7 | Limiters - Publication Date: -20241231 | N/A | 1881 |
| 8 | Limit to English | 1480 | 1871 |
| 9 | Advanced Search function to exclude Medline records | **418** | 611 |
| 10 | Limiters - Publication Date: 2021/02/01-2024/12/31 | N/A | **219** |

**Supplemental File B:** Detailed Voting Instructions for the Three-Round Delphi Consultation

**Delphi Round 1**:

Stakeholders were sent a personalised link to an online Excel document, named according to their pseudonymous participant ID, containing the acute audit items (*n*=200 items in total: *n*=103 INAS items and *n*=97 frequently occurring international items). The items were listed alongside the frequency counts displaying the number of other international stroke audits and registries, aside from the audit that the item was derived from, that collected that datapoint. The frequency counts were also colour-coded according to a traffic-light system to aid interpretation:

- ‘Green’ represented that the index or comparator item, namely the audit item being cross-checked, was shared by a high number of audits/registries, defined as 12-21 audits/registries in *total* or 11-20 audits/registries *excluding* the comparator audit.
- ‘Yellow’ showed that the index item was shared with a moderate number of other audits/registries (i.e., 4-11 audits/registries in *total* or 3-10 audits/registries *excluding* the comparator audit).
- For INAS items only, ‘dark orange’ showed that the Irish audit item was either not shared, or shared with very few, other registries (i.e., 0-3 audits/registries in *total* or 0-2 audits/registries *excluding* the comparator INAS audit item). International items occurring in fewer than 4 audits in total were excluded from review.

For each item, stakeholders were asked to select from the following response options: “Include in INAS”, “Exclude from INAS”, “No strong view”, or “Don’t know enough to be able to say”. Participants were also provided with space to leave any qualitative feedback regarding each item, if desired. A two-week deadline was provided to respond to this task, with follow-up of stakeholders who did not respond within this timeframe.

The breakdown of the percentage of participating stakeholders who voted (anonymously) for each response option and a summary of the anonymous qualitative item feedback were aggregated and presented at both project Steering Group and INAS Governance Committee meetings. The descriptive results of Round 1 votes were circulated to participants in tabular form.

**Delphi Round 2**:

Initially responding stakeholders (i.e., those participants who responded to Round 1) were sent a task document that showed their responses to Round 1 alongside the mean aggregated group responses for each item. Participants were provided with an opportunity to revise their original response based on this comparative feedback, if desired, and submit amended responses within two weeks. Participants were notified that if they did not respond within the two-week timeframe, it would be assumed that they were happy with their previous responses and did not wish to make any changes.

The results of the second round of the Delphi consultation were categorised according to consensus criteria by the mean percentage of votes to include the item as part of INAS. Specifically, Irish and international acute stroke care items were each classified as receiving a high (≥70%), moderate (50-69%), or low (<50%) percentage of votes to include in INAS. These descriptive results, as well as any open-ended qualitative responses, were presented and discussed at subsequent stakeholder meetings. Additional feedback generated from the meeting discussions was transcribed and an anonymised summary of the feedback was incorporated into the qualitative notes from previous Delphi rounds in an iterative cycle of controlled feedback. Based on stakeholder engagement, additional items were proposed during this stage and added to the inventory of audit items for consideration in Round 3 ($n$ = 22).

As per the consensus reached at the Steering Group and Governance Committee meetings, the low-voted INAS Core Clinical items and low-voted international items were excluded from consideration. The highly-voted INAS HIPE, Core Clinical, and Thrombectomy items were recommended for ongoing inclusion in INAS, with no further discussion in Round 3 required. All remaining items (i.e., the moderately-voted INAS Core Clinical items, low-voted INAS Thrombectomy items, highly-voted international items, moderately voted international items, and those newly proposed items derived from stakeholder engagement) were put forward to a third, and final, Delphi round.

**Delphi Round 3:**

For a reduced set of items, all eligible stakeholders were asked for a final time to vote “Include in INAS”, “Exclude from INAS”, or “Don’t know enough to be able to say”. Stakeholders were provided with an anonymised summary of the discussion points arising from all previous meetings to help inform their decisions. A two-week deadline was given for task completion.

The third Delphi round item group votes were tallied and classified as receiving high, moderate, or low mean percentages to include as part of INAS, as before. These results were presented and discussed at additional project Steering Group and INAS Governance Committee meetings. These meetings provided a final opportunity for the expert members to provide any clinical or policy justifications for the removal or retention of items and to provide final consensus to deliver the core minimum dataset for audit of acute stroke care in Ireland.

**Supplemental File C.** Characteristics of included national and continuous stroke audits and registries.

| Country | Registry Name | Year Started | Participating Hospitals | Number Registered in Last Annual Report | Types of Cases Included | Type of Care Included | Coverage | Participation |
|---|---|---|---|---|---|---|---|---|
| **Australia** | **Australian Stroke Clinical Registry (AuSCR)** | 2009 | 61 participating hospitals | 17184 in 2022 | Ischaemic stroke; Haemorrhagic stroke; TIA | Acute hospital care; Follow-up; Stroke Foundation conducts organisational survey and rehab on a subsection of patients | Not specified | Voluntary |
| **Austria** | **The Austrian Stroke Unit Registry** | 2003 | All 38 stroke units report | Not specified | Ischaemic stroke; Haemorrhagic stroke; SAH | Risk factors; Acute hospital Care; Follow-up | 2/3 of patients with stroke in Austria (1/3 are treated on other wards) | Mandated |
| **Canada** | **CorHealth Ontario** | Previously: Registry of the Canadian Stroke Network; CorHealth Ontario from 2017 | Not specified | Not specified | Ischaemic stroke; Haemorrhagic stroke; TIA; SAH | Acute hospital care; Post-acute rehabilitation; Follow-up | Not specified | Not specified |
| **Denmark** | **DanStroke** | 2003 | All hospitals treating patients with stroke report | 14853 | Ischaemic stroke; Haemorrhagic stroke; TIA; SAH | Risk factors; Acute hospital Care; Follow-up | >90% | Mandated |

| | | | | | | | | |
|---|---|---|---|---|---|---|---|---|
| **Germany** | **German Stroke Register Study Group** | 1999 | 736 hospitals in 2016 (A network of 10 regional quality assurance projects) | Approximately 300 000 each year | Ischaemic stroke; Haemorrhagic stroke; TIA; SAH; Separate thrombectomy registry | Risk factors; Acute hospital Care | Not specified | Mandated for all stroke units and all hospitals in some regions |
| **Ireland** | **Irish National Audit of Stroke** | 2013 as National Stroke Register | 21 Hospitals All hospitals with >25 stroke admissions/year | 4999 in 2022 | Ischaemic stroke; Haemorrhagic stroke | Acute hospital Care; Organisational audit | 90% | Voluntary |
| **Japan** | **The Japan Stroke Data Bank (previously the Japan Standard Stroke Registry Study)** | 1999 | 132 hospitals participate as of 2021 | 19841 in 2020 | Ischaemic stroke; Haemorrhagic stroke; TIA; SAH | Risk factors; Acute hospital care; Follow-up | Approximately 6% | Not specified |
| **Netherlands** | **Dutch Acute Stroke Audit** | 2014 | Not specified | Approximately 30000 annually | Ischaemic stroke; Haemorrhagic stroke; TIA | Acute hospital care Follow-up | Not specified | Voluntary |
| **Norway** | **Norwegian Stroke Registry (NHR)** | 2012 | All hospitals involved in stroke care (51 hospitals) | 9020 in 2022 | Ischaemic stroke; Haemorrhagic stroke; | Risk factors; Pre-hospital care; Acute hospital Care; Follow-up | 89% | Mandated |
| **Scotland** | **Scottish Stroke Care Audit (SSCA)** | 2002 | All hospitals providing stroke care | 11257 in 2022 | Ischaemic stroke; Haemorrhagic stroke; TIA; Separate thrombectomy audit | Pre-hospital; Acute hospital care; Follow-up | Not specified | Mandated |
| **Singapore** | **Singapore Stroke Registry** | 2002 | All public healthcare institutions | 9680 in 2021 | Ischaemic stroke; Haemorrhagic stroke; SAH | Risk factors; Acute hospital care; Follow-up | Approximately 94% of stroke patients | Not specified |

| | | | | | | | | |
|---|---|---|---|---|---|---|---|---|
| **Slovakia** | **Slovak Stroke Register** | 2010 | All 43 stroke centres report | Not specified | Ischaemic stroke; Haemorrhagic stroke; TIA; SAH | Risk factors; Acute hospital care | Not specified | Mandated |
| **South Korea** | **Korean Stroke Registry (KSR)** | 2001 | 76 Hospitals | 21789 in 2021 | Ischaemic stroke; Haemorrhagic stroke; TIA | Risk factors; Acute hospital care | Not specified | Voluntary |
| **South Korea** | **CRCS-K DB (Clinical Research Collaboration for Stroke in Korea)** | 2008 | 17 university hospitals or regional stroke centres | 8057 in 2019 | Ischaemic stroke; TIA | Risk factors; Acute hospital care; Follow-up | 10.7% of AIS patients in South Korea | Not specified |
| **Spain** | **RENISEN** | 2011 | 42 primary stroke centers in Spain in 2016 | Not specified | Ischaemic stroke; Haemorrhagic stroke; TIA | Risk factors; Acute hospital care; Follow-up | Not specified | Not specified |
| **Sweden** | **Riksstroke** | 1994 | All 72 hospitals treating acute stroke | 20115 Strokes and 8797 TIAs in 2022 | Ischaemic stroke; Haemorrhagic stroke; TIA; SAH | Risk factors; Pre-hospital care; Acute hospital care; Post-acute rehabilitation; Follow-up | Approximately 95% | Mandated |
| **Switzerland** | **Swiss Stroke Registry** | 2014 | All hospitals certified in stroke therapy, 10 Stroke Centres and 16 Stroke Units or network hospitals | >10000 registered annually | Ischaemic stroke; Haemorrhagic stroke | Risk factors; Acute hospital care; Follow-up | Not specified | Mandated |
| **Taiwan** | **Taiwan Stroke Registry** | 2006 | 65 hospitals | Not specified | Ischaemic stroke; Haemorrhagic stroke; TIA; SAH | Risk factors; Acute hospital care; Follow-up | Approximately 18% | Not specified |

| | | | | | | | | |
|---|---|---|---|---|---|---|---|---|
| **UK** | **Sentinel Stroke National Audit Programme** | 1998 (Under other names initially) | All hospitals providing stroke care | 91,162 in 2023 report | Ischaemic stroke<br>Haemorrhagic stroke<br>TIA | Risk factors;<br>Pre-hospital care;<br>Acute hospital care;<br>Post-acute rehabilitation;<br>Organisational audit;<br>Follow-up | >90% | Mandated |
| **United States** | **Paul Coverdell National Acute Stroke Program** | 2001 | 13 states are recipients of funding | Not specified | Ischaemic stroke;<br>Haemorrhagic stroke; TIA;<br>SAH | Risk factors;<br>Pre-hospital care;<br>Acute hospital care;<br>Follow-up | Not specified | Voluntary with financial incentive |
| **United States** | **Get with the Guidelines Stroke Registry** | 2003 | >2000 hospitals participate covering >50% of hospitals that admit strokes in US | Not specified | Ischaemic stroke;<br>Haemorrhagic stroke; TIA: SAH | Risk factors;<br>Pre-hospital;<br>Acute hospital care;<br>Follow-up | Not specified | Voluntary |

***Note***. Abbreviations: TIA: Transient Ischaemic Attack; SAH: Subarachnoid haemorrhage.

**Supplemental File D.** Phase 1 Benchmarking: Comparison of INAS Core Clinical and Thrombectomy Items against International Stroke Audits and Registries

| | | Legend |
|---|---|---|
| Low | 0 | INAS item is not shared by any other audit/registry |
| | 1-2 | INAS item is shared by 1-2 other audits/registries (or 2-3 total) |
| Moderate | 3-5 | INAS item is shared by 3-5 other audits/registries (or 4-6 total) |
| | 6-10 | INAS item is shared by 6-10 other audits/registries (or 7-11 total) |
| High | 11-15 | INAS item is shared by 11-15 other audits/registries (or 12-16 total) |
| | 16-20 | INAS item is shared by 16-20 other audits/registries (or 17-21 total) |

| Irish Audit (INAS) Dataset | INAS Audit Item Descriptor | Number of *Other* International Audits/Registries Collecting Similar or Closely Related Items | | |
|---|---|---|---|---|
| | | Same Item | Related Item | Combined (Same + Related Items) |
| HIPE Dataset | Age | 9 | 0 | **9** |
| HIPE Dataset | Date of discharge | 14 | 1 | **15** |
| HIPE Dataset | Hospital acquired diagnosis (1-30) (diagnosis acquired during the patient's episode of care that was not present on admission, 1st additional diagnosis (DIAG2).) - if flagged as hospital acquired diagnosis of stroke | 2 | 3 | **5** |
| HIPE Dataset | Length of stay (For in-patient discharges, this is the number of days between the admission date and discharge date, not including the final day (discharge date). It is also sometimes referred to as Bed Days.) | 5 | 3 | **8** |
| HIPE Dataset | Principal diagnosis (The diagnosis established after study to be chiefly responsible for occasioning the episode of admitted patient care.) - 8th ed ICD-10-AM/ACHI/ACS and ICS | 17 | 3 | **20** |
| HIPE Dataset | Sex | 17 | 0 | **17** |
| HIPE Dataset | Source of admission (Describes where the patient was admitted from. It does not refer to where an emergency or accident occurred.) – 01 home, 02 transfer from nursing home, convalescent home or other long-stay accommodation, 03 transfer of admitted patient from acute hospital, 04 transfer from non-acute Hospital, 05 transfer from hospice, 06 | 5 | 4 | **9** |

| | | | | |
|---|---|---|---|---|
| | transfer from psychiatric hospital/unit, 07 new born, 08 temporary place of residence, 09 prison, 0 other | | | |
| HIPE Dataset | Type of admission (The category of admission relating to this episode of care, indicates the priority of the admission.) – 1 elective, 2 elective readmission, 4 emergency, 5 emergency readmission, 6 maternity, 7 new-born | 0 | 3 | **3** |
| HIPE Dataset | 1st additional diagnosis (A condition or complaint either coexisting with the principal diagnosis or arising during the episode of admitted patient care. Interpreted as conditions that affect patient management.) | 1 | 5 | **6** |
| HIPE Dataset | Hospital code list | 9 | 3 | **12** |
| Core Clinical | Which hospital was patient transferred from (if any) | 8 | 5 | **13** |
| Core Clinical | Why was the patient transferred - 1 Thrombolysis 2 Thrombectomy 3 Neurosurgery 8 Other | 3 | 1 | **4** |
| Core Clinical | If other transfer reason, please specify | 1 | 0 | **1** |
| Core Clinical | If other transfer hospital, please specify | 0 | 0 | **0** |
| Core Clinical | Symptom onset date | 19 | 1 | **20** |
| Core Clinical | Symptom onset time | 19 | 1 | **20** |
| Core Clinical | If symptom onset date is unknown, what date was the patient last known to be well | 9 | 0 | **9** |
| Core Clinical | If symptom onset time is unknown, what time was the patient last known to be well | 11 | 0 | **11** |
| Core Clinical | Did the stroke occur while the patient was in hospital for treatment of another condition - 1 Yes 2 No 9 Unknown | 9 | 2 | **11** |
| Core Clinical | If no, date of presentation to hospital | 16 | 2 | **18** |
| Core Clinical | If no, time of presentation to hospital | 16 | 2 | **18** |
| Core Clinical | If presentation time is unknown, was presentation to hospital within 4.5 hrs of symptom onset - 1 Yes, 2 No 9 Unknown | 0 | 5 | **5** |
| Core Clinical | Medical assessment date | 2 | 1 | **3** |
| Core Clinical | Medical assessment time | 4 | 2 | **6** |
| Core Clinical | Brain CT or MRI performed - 1 Yes, 2 No 3 Performed pre-admission / hospital transfer 9 Unknown | 17 | 1 | **18** |

| | | | | |
|---|---|---|---|---|
| Core Clinical | If yes, First Brain Imaging date | 16 | 1 | **17** |
| Core Clinical | If yes, First Brain Imaging time | 16 | 0 | **16** |
| Core Clinical | Did the patient receive I.V. Thrombolysis - 1 Yes, 2 No 5 Contraindicated | 17 | 2 | **19** |
| Core Clinical | If yes, enter date | 18 | 0 | **18** |
| Core Clinical | If yes, enter time | 18 | 0 | **18** |
| Core Clinical | If yes, was intracerebral bleed seen on scan within 36 hrs - 1 Yes, 2 No 9 Unknown | 6 | 7 | **13** |
| Core Clinical | If intracerebral bleed, was neuro deterioration associated with it - 1 Yes, 2 No 9 Unknown | 0 | 10 | **10** |
| Core Clinical | Did the patient have thrombectomy in this hospital (Beaumont / CUH only) - 1 Yes, 2 No | 8 | 7 | **15** |
| Core Clinical | Was a swallow screen completed - 1 Yes, 2 No 9 Unknown | 14 | 0 | **14** |
| Core Clinical | If yes, was swallow screen completed within 4 hours of presentation - 1 Yes, 2 No 9 Unknown | 2 | 5 | **7** |
| Core Clinical | Modified Rankin Scale pre-stroke - 0 Zero 1 One 2 Two 3 Three 4 Four 5 Five 6 Six 9 Unknown | 10 | 3 | **13** |
| Core Clinical | Admitted to Stroke Unit (Key Performance Indicator) - 1 Yes, 2 No | 13 | 1 | **14** |
| Core Clinical | If yes, date admitted to Stroke Unit (Key Performance Indicator) | 6 | 1 | **7** |
| Core Clinical | If yes, date discharged from Stroke Unit (Key Performance Indicator) | 5 | 0 | **5** |
| Core Clinical | If no, reason why - 1 No Stroke Unit 2 Bed Not Available 5 Infection Control Risk 8 Other | 1 | 1 | **2** |
| Core Clinical | If other reason, please specify | 0 | 0 | **0** |
| Core Clinical | Allied Health Professional (AHP) Assessment - 1 Yes, 2 No | 4 | 6 | **10** |
| Core Clinical | If yes, Physiotherapy - 1 Yes, 2 No 3 Not Indicated 9 Unknown | 6 | 2 | **8** |
| Core Clinical | If yes, Occupational Therapy - 1 Yes, 2 No 3 Not Indicated 9 Unknown | 5 | 2 | **7** |
| Core Clinical | If yes, Speech and Language Therapy - 1 Yes, 2 No 3 Not Indicated 9 Unknown | 5 | 2 | **7** |
| Core Clinical | If yes, Dietetics - 1 Yes, 2 No 3 Not Indicated 9 Unknown | 3 | 1 | **4** |

| | | | | |
|---|---|---|---|---|
| Core Clinical | If yes, Medical Social Worker - 1 Yes, 2 No 3 Not Indicated 9 Unknown | 1 | 2 | **3** |
| Core Clinical | If yes, Psychology - 1 Yes, 2 No 3 Not Indicated 9 Unknown | 1 | 1 | **2** |
| Core Clinical | Was the patient assessed by Stroke Nurse Specialist - 1 Yes, 2 No 3 Not Indicated 9 Unknown | 1 | 2 | **3** |
| Core Clinical | If no, reason why | 1 | 0 | **1** |
| Core Clinical | Multidisciplinary Meeting Case assessment -1 Yes, 2 No 3 Not Indicated 9 Unknown | 1 | 1 | **2** |
| Core Clinical | Was an assessment of mood completed and documented by a member of the multidisciplinary team - 1 Yes, 2 No 3 Not Indicated 9 Unknown | 4 | 0 | **4** |
| Core Clinical | Does the patient have Symptomatic Carotid Stenosis -1 Yes, 2 No 9 Unknown | 3 | 3 | **6** |
| Core Clinical | If Symptomatic Carotid Stenosis, was the patient referred for Carotid Endarterectomy - 1 Yes, 2 No 9 Unknown | 5 | 1 | **6** |
| Core Clinical | If Symptomatic Carotid Stenosis, was the patient referred for Carotid Stenting - 1 Yes, 2 No 9 Unknown | 6 | 1 | **7** |
| Core Clinical | New or Altered Antithrombotic Therapy prescribed for acute treatment - 1 Yes, 2 No 3 Contraindicated 9 Unknown | 11 | 4 | **15** |
| Core Clinical | If yes, Antiplatelet or Anticoagulant (for acute treatment) start date | 3 | 4 | **7** |
| Core Clinical | Does the patient have Atrial Fibrillation - 1 Yes, 2 No 4 Results Pending 9 Unknown | 15 | 2 | **17** |
| Core Clinical | If Atrial Fibrillation, was atrial fibrillation known prior to stroke onset - 1 Yes, 2 No 9 Unknown | 12 | 3 | **15** |
| Core Clinical | If atrial fibrillation known prior to stroke onset, was Antiplatelet And/Or Anticoagulant prescribed prior to Stroke onset - 1 Yes, 2 No 9 Unknown | 12 | 1 | **13** |
| Core Clinical | If yes, please specify Antiplatelet / Anticoagulant - prior to stroke – 0 NOAC 1 Warfarin 5 Aspirin 6 Clopidogrel 7 Other Antiplatelet 8 Dual Antiplatelet Therapy 9 Antiplatelet & Anticoagulant | 10 | 2 | **12** |
| Core Clinical | 1 Yes, 2 No 9 Unknown If atrial fibrillation known prior to stroke onset, and on Warfarin, was INR (International Normalised Ratio) 2-3 at Stroke onset. | 6 | 3 | **9** |
| Core Clinical | If Atrial Fibrillation, Anticoagulation prescribed for secondary prevention - 1 Yes, 2 No 9 Unknown | 12 | 5 | **17** |
| Core Clinical | If yes, please specify Antiplatelet / Anticoagulant - on discharge – 0 NOAC 1 Warfarin 5 Aspirin 6 Clopidogrel 7 Other Antiplatelet 8 Dual Antiplatelet Therapy 9 Antiplatelet & Anticoagulant | 12 | 1 | **13** |

| | | | | |
|---|---|---|---|---|
| Core Clinical | If no, please enter reason documented - 01 No reason documented 02 Major bleeding (prior history) 03 Severe illness (e.g. cancer, dementia) 04 Poor compliance (known or suspected) 05 Patient refused anticoagulation 06 Alcohol excess 07 Falls 08 Extreme frailty 09 Liver disease | 5 | 1 | **6** |
| Core Clinical | Modified Rankin Scale on discharge - 0 Zero 1 One 2 Two 3 Three 4 Four 5 Five 6 Six 9 Unknown | 14 | 0 | **14** |
| Core Clinical | Discharge destination - 1 Home 2 Patient died 3 Discharge to long term care 4 Discharge to off-site rehab 5 Transfer to referring hospital 6 Transfer to other hospital for on-going stroke care 7 Home with ESD 8 Other 9 Unknown | 14 | 2 | **16** |
| Core Clinical | Case complete - 1 Yes, 2 No, 9 Unknown | 0 | 2 | **2** |
| Thrombectomy | NIHSS pre-thrombectomy | 5 | 14 | **19** |
| Thrombectomy | Date of performance of non-contrast CT | 5 | 3 | **8** |
| Thrombectomy | Time of performance of non-contrast CT | 4 | 2 | **6** |
| Thrombectomy | Date of performance of non-contrast CTA | 7 | 6 | **13** |
| Thrombectomy | Time of performance of non-contrast CTA | 4 | 4 | **8** |
| Thrombectomy | Date of contact with the endovascular stroke centre | 1 | 1 | **2** |
| Thrombectomy | Time of contact with the endovascular stroke centre | 1 | 0 | **1** |
| Thrombectomy | Date of decision to transfer patient | 2 | 4 | **6** |
| Thrombectomy | Time of decision to transfer patient | 2 | 1 | **3** |
| Thrombectomy | Was the patient transferred from another hospital - 1 Yes, 2 No 9 Unknown | 9 | 2 | **11** |
| Thrombectomy | If yes, what date did the patient arrive at the referring/first hospital | 17 | 1 | **18** |
| Thrombectomy | If yes, what time did the patient arrive at the referring/first hospital | 17 | 1 | **18** |
| Thrombectomy | If yes, what date did the patient leave the referring/first hospital for transfer to the EVT centre? | 4 | 1 | **5** |
| Thrombectomy | If yes, what time did the patient leave the referring/first hospital for transfer to the EVT centre? | 3 | 1 | **4** |

| | | | | |
|---|---|---|---|---|
| Thrombectomy | Date of arrival at the endovascular stroke centre | 3 | 3 | **6** |
| Thrombectomy | Time of arrival at the endovascular stroke centre | 3 | 3 | **6** |
| Thrombectomy | Did the patient have repeat non-invasive imaging in the endovascular stroke centre - 1 Yes, 2 No 9 Unknown | 2 | 4 | **6** |
| Thrombectomy | If yes, please specify - 1 Non-contrast CT 2 CTA 3 Perfusion CT 4 MRI | 1 | 3 | **4** |
| Thrombectomy | Site of most proximal occlusion - 1 MCA 1 2 MCA 2 3 Basilar 4 ICA carotid T/L 5 ICA cervical segment 6 PCA 7 Vertebro basilar | 8 | 3 | **11** |
| Thrombectomy | Second occlusion site | 1 | 1 | **2** |
| Thrombectomy | Associated carotid stenosis greater than 50% - 1 Yes, 2 No 9 Unknown | 7 | 1 | **8** |
| Thrombectomy | TICI pre thrombectomy | 4 | 2 | **6** |
| Thrombectomy | TICI post thrombectomy | 10 | 1 | **11** |
| Thrombectomy | Date of groin puncture | 9 | 1 | **10** |
| Thrombectomy | Time of groin puncture | 10 | 1 | **11** |
| Thrombectomy | Date of first pass | 1 | 3 | **4** |
| Thrombectomy | Time of first pass | 1 | 1 | **2** |
| Thrombectomy | Date of first reperfusion | 7 | 3 | **10** |
| Thrombectomy | Time of first reperfusion | 6 | 3 | **9** |
| Thrombectomy | Date of final angio | 0 | 2 | **2** |
| Thrombectomy | Time of final angio | 0 | 2 | **2** |
| Thrombectomy | Immediate complications - Not Applicable 1 Haemorrhage 2 Embolus into separate vascular territory 3 Dissection 8 Other 9 Unknown | 5 | 3 | **8** |
| Thrombectomy | NIHSS 24-hour post-thrombectomy | 9 | 5 | **14** |

| | | | | |
|---|---|---|---|---|
| Thrombectomy | Following procedure was patient transferred immediately back to primary receiving hospital - 1 Yes, 2 No 9 Unknown | 2 | 3 | **5** |
| Thrombectomy | If no, when was patient admitted to the endovascular stroke centre - 1 0-3 hrs 2 3-12 hrs 3 12-24 hrs 4 24+ hrs | 0 | 1 | **1** |

***Note.*** Each row contains information on the INAS dataset that the data item pertains to and the audit item descriptor. In Phase 1 of the benchmarking exercise, the items from the HIPE and INAS Core Clinical and Thrombectomy datasets were cross-checked against 20 international audits and registries to see how frequently other registries ask something equivalent or related. The benchmarking frequency counts are available for each item in the table. Note that these frequency counts do not include the Irish INAS index item itself within the count; hence, where the count says 3 that means 3 other audits/registries share the item in addition to INAS. The items are colour-coded based on a traffic light system; whereby, dark green represents that the item is shared by a high number of registries, yellow marks that the item is shared with a moderate number of other registries, and dark orange meaning the item is not shared by any other registry.

Abbreviations: Angio: angiogram; CT: computed tomography; CTA: computed tomography angiography; CUH: Cork University Hospital; ESD: early supported discharge; EVT: endovascular thrombectomy; HIPE: Hospital In-Patient Enquiry; ICA: internal carotid artery; IV: intravenous; MCA: middle cerebral artery; NIHSS: National Institutes of Health Stroke Scale; NOAC: non-vitamin K oral anticoagulants; PCA: posterior cerebral artery; TICI: thrombolysis in cerebral infarction.

**Supplemental File E.** Phase 2 Benchmarking: Frequency Counts of International Acute Stroke Care Audit Items, not collected by INAS

| | Legend | |
|---|---|---|
| Moderate | 3-5 | International reference item is shared by 3-5 other audits/registries (or 4-6 total) |
| | 6-10 | International reference item is shared by 6-10 other audits/registries (or 7-11 total) |
| High | 11-15 | International reference item is shared by 11-15 other audits/registries (or 12-16 total) |
| | 16-19 | International reference item is shared by 16-19 other audits/registries (or 17-20 total) |

| Country of Audit | International Audit Reference Item Descriptor | Number of *Other* International Audits/Registries Collecting Similar or Closely Related Items | | |
|---|---|---|---|---|
| | | Same Item | Related Item | Combined (Same + Related Items) |
| Australia AuSCR | Patient Record ID [Patient Details] | 6 | 4 | **10** |
| Australia AuSCR | First name [Patient Details] | 9 | 0 | **9** |
| Australia AuSCR | Last name [Patient Details] | 10 | 0 | **10** |
| Australia AuSCR | Country of birth [Patient Details] | 3 | 0 | **3** |
| Australia AuSCR | Postcode [Contact Information] | 8 | 0 | **8** |
| Australia AuSCR | Transport code [Pre-Hospital Care] | 0 | 3 | **3** |
| Australia AuSCR | Did the patient arrive by ambulance? [Admission Details] | 6 | 0 | **6** |
| Australia AuSCR | Previous stroke (A history of stroke prior to this current episode, excluding TIAs) [History of Known Risk Factors] | 14 | 0 | **14** |
| Australia AuSCR | Previous TIA (Transient Ischemic Attack (TIA) or mini stroke) [History of Known Risk Factors] | 12 | 0 | **12** |
| Australia AuSCR | Diabetes [History of Known Risk Factors] | 16 | 0 | **16** |
| Australia AuSCR | Hypercholesterolaemia [History of Known Risk Factors] | 8 | 0 | **8** |

| | | | | |
|---|---|---|---|---|
| Australia AuSCR | Hypertension [History of Known Risk Factors] | 14 | 0 | **14** |
| Australia AuSCR | Recent myocardial infarction [History of Known Risk Factors] | 8 | 0 | **8** |
| Australia AuSCR | Ischaemic heart disease [History of Known Risk Factors] | 7 | 0 | **7** |
| Australia AuSCR | High alcohol consumption [History of Known Risk Factors] | 8 | 0 | **8** |
| Australia AuSCR | Current smoker [History of Known Risk Factors] | 14 | 0 | **14** |
| Australia AuSCR | Past smoker [History of Known Risk Factors] | 13 | 0 | **13** |
| Australia AuSCR | Family history of stroke [History of Known Risk Factors] | 4 | 0 | **4** |
| Australia AuSCR | Obesity [History of Known Risk Factors] | 3 | 0 | **3** |
| Australia AuSCR | Peripheral vascular disease [History of Known Risk Factors] | 7 | 0 | **7** |
| Australia AuSCR | Other serious illness that influences prognosis or management of stroke [History of Known Risk Factors] | 3 | 2 | **5** |
| Australia AuSCR | Did the patient take antihypertensives prior to this stroke? [Pre-Admission Medication] | 7 | 0 | **7** |
| Australia AuSCR | Did the patient take lipid lowering treatment prior to this stroke? [Pre-Admission Medication] | 5 | 0 | **5** |
| Australia AuSCR | Statin [Pre-Admission Medication] | 4 | 0 | **4** |
| Australia AuSCR | Can the patient walk on their own (i.e. without the assistance of another person, but may include walking aid)? - Pre-stroke functional status. [Dependency Prior to Admission] | 2 | 1 | **3** |
| Australia AuSCR | Living arrangements prior to admission [Dependency Prior to Admission] | 6 | 0 | **6** |
| Australia AuSCR | Assessment for level of responsiveness [Acute Clinical Data] | 2 | 1 | **3** |
| Australia AuSCR | Was the patient unresponsive (or in a coma)? [Acute Clinical Data] | 2 | 1 | **3** |
| Australia AuSCR | Patient’s glucose level before thrombolysis treatment or on hospital arrival (if no thrombolysis) amount (mmol/l) [Acute Clinical Data] | 9 | 0 | **9** |
| Australia AuSCR | NIHSS at baseline [Acute Clinical Data] | 6 | 1 | **7** |
| Australia AuSCR | Weight at baseline, Amount (kg) [Acute Clinical Data] | 6 | 0 | **6** |

| | | | | |
|---|---|---|---|---|
| Australia AuSCR | BP at baseline (SBP/DBP) [Acute Clinical Data] | 11 | 0 | **11** |
| Australia AuSCR | What type of brain scan was performed? [Brain Imaging] | 3 | 0 | **3** |
| Australia AuSCR | Type of stroke [Brain Imaging] | 2 | 1 | **3** |
| Australia AuSCR | Side of stroke affected [Brain Imaging] | 3 | 0 | **3** |
| Australia AuSCR | Cause of stroke (Stroke cause determined based on TOAST classification system) [Brain Imaging] | 4 | 0 | **4** |
| Australia AuSCR | Mechanism (The cause or mechanism of current stroke episode.) [Brain Imaging] | 5 | 0 | **5** |
| Australia AuSCR | Did the patient have a 12 lead ECG in hospital? [Other Investigations] | 6 | 2 | **8** |
| Australia AuSCR | Type of other cardiac investigations - If yes, specify [Other Investigations] | 3 | 0 | **3** |
| Australia AuSCR | Drug (Drug used for intravenous thrombolysis) [Telemedicine Setting and Reason] | 4 | 0 | **4** |
| Australia AuSCR | Dose (Full (0.9mg/kg)) (The total number of milligrams of intravenous thrombolysis therapy given intravenously (combined amount of bolus and infusion amount)) [Telemedicine Setting and Reason] | 4 | 0 | **4** |
| Australia AuSCR | Was patient outside of time window? (Patient was outside of the window of time when thrombolysis can safely be given) [Telemedicine Setting and Reason] | 2 | 1 | **3** |
| Australia AuSCR | Patient was fully worked up within 4.5 hours but tPA unable to be given for following reasons [Telemedicine Setting and Reason] | 2 | 2 | **4** |
| Australia AuSCR | Age> 80 (Reason(s) why thrombolysis was not given (despite patient being fully worked up within 4.5 hours).) [Telemedicine Setting and Reason] | 3 | 0 | **3** |
| Australia AuSCR | Major co-morbidity (Reason(s) why thrombolysis was not given (despite patient being fully worked up within 4.5 hours).) [Telemedicine Setting and Reason] | 3 | 0 | **3** |
| Australia AuSCR | Warfarin with INR> 1.7 (Reason(s) why thrombolysis was not given (despite patient being fully worked up within 4.5 hours).) [Telemedicine Setting and Reason] | 1 | 2 | **3** |
| Australia AuSCR | Rapidly improving (Reason(s) why thrombolysis was not given (despite patient being fully worked up within 4.5 hours).) [Telemedicine Setting and Reason] | 5 | 0 | **5** |
| Australia AuSCR | Other contraindication (Reason(s) why thrombolysis was not given (despite patient being fully worked up within 4.5 hours).) [Telemedicine Setting and Reason] | 5 | 0 | **5** |
| Australia AuSCR | Anaesthesia used (Type of anaesthesia used during reperfusion treatment) [Telemedicine Setting and Reason] | 4 | 0 | **4** |
| Australia AuSCR | Primary/Initial IA treatment (Primary thrombectomy method/device used) [Telemedicine Setting and Reason] | 3 | 1 | **4** |
| Australia AuSCR | Heparin (Heparin used during endovascular treatment.) [Telemedicine Setting and Reason] | 3 | 0 | **3** |
| Australia AuSCR | If yes, were fluids administered via IV or nasogastric tube? [Hydration and Nutrition] | 2 | 1 | **3** |

| | | | | |
|---|---|---|---|---|
| Australia AuSCR | Did the patient with intracerebral haemorrhage receive hemicraniectomy [Surgical Management] | 5 | 3 | **8** |
| Australia AuSCR | Date of hemicraniectomy [Surgical Management] | 3 | 0 | **3** |
| Australia AuSCR | Was a venous blood glucose level sample collected and sent to laboratory while patient was in the ED? [Assessment and Management of Hyperglycaemia] | 3 | 3 | **6** |
| Australia AuSCR | Did management of the patient include Heparin (or Low Molecular Weight Heparin)? [DVT Prophylaxis] | 5 | 0 | **5** |
| Australia AuSCR | Aspiration pneumonia [Complications during Hospital Admission] | 6 | 0 | **6** |
| Australia AuSCR | Deep Vein Thrombosis (DVT) [Complications during Hospital Admission] | 6 | 0 | **6** |
| Australia AuSCR | Pulmonary Thrombo-Embolism (PE) [Complications during Hospital Admission] | 3 | 0 | **3** |
| Australia AuSCR | Acute myocardial infarction [Complications during Hospital Admission] | 3 | 0 | **3** |
| Australia AuSCR | New stroke [Complications during Hospital Admission] | 5 | 0 | **5** |
| Australia AuSCR | Urinary tract infection [Complications during Hospital Admission] | 4 | 0 | **4** |
| Australia AuSCR | Seizures [Complications during Hospital Admission] | 4 | 0 | **4** |
| Australia AuSCR | Other [Complications during Hospital Admission] | 3 | 0 | **3** |
| Australia AuSCR | On discharge was the patient prescribed antihypertensive agents? [Medication Prescribed at Discharge] | 5 | 0 | **5** |
| Australia AuSCR | On discharge was the patient prescribed lipid lowering treatment? [Medication Prescribed at Discharge] | 5 | 1 | **6** |
| Australia AuSCR | Statin [Medication Prescribed at Discharge] | 9 | 0 | **9** |
| Australia AuSCR | Patient deceased during hospital care? [Discharge Information] | 3 | 1 | **4** |
| Australia AuSCR | Date of death [Discharge Information] | 5 | 0 | **5** |
| Australia AuSCR | Cause of death [Discharge Information] | 5 | 1 | **6** |
| Australia AuSCR | NIHSS at discharge [Dependency on Discharge] | 3 | 0 | **3** |
| Australia AuSCR | Date of NIHSS if not at discharge (e.g. day 2-7) [Dependency on Discharge] | 3 | 0 | **3** |

| | | | | |
|---|---|---|---|---|
| Australia AuSCR | Death date post-acute episode of care [Record Death for Patient] | 1 | 3 | **4** |
| Sweden RIKSSTROKE | Enter hospital code where thrombolysis was performed [Thrombolysis] | 2 | 1 | **3** |
| South Korea KSR | Acute Treatment: door to needle time | 5 | 0 | **5** |
| UK SSNAP | Was the patient given anticoagulant reversal therapy? | 1 | 2 | **3** |
| South Korea CRCS-5 | Study ID | 3 | 0 | **3** |
| South Korea KSR | Stroke subtype: Haemorrhage-ICH | 2 | 1 | **3** |
| South Korea KSR | Stroke subtype: Haemorrhage-SAH | 0 | 4 | **4** |
| South Korea KSR | Stroke subtype: TIA | 1 | 3 | **4** |
| South Korea KSR | Acute Treatment: other reasons for no IV tPA - text | 0 | 4 | **4** |
| South Korea KSR | Vital/physical: height | 5 | 0 | **5** |
| South Korea KSR | Laboratory test: Hb | 3 | 0 | **3** |
| South Korea KSR | Laboratory test: Total Chol | 7 | 0 | **7** |
| South Korea KSR | Laboratory test: HDL Chol | 6 | 0 | **6** |
| South Korea KSR | Laboratory test: TG | 3 | 0 | **3** |
| South Korea KSR | Laboratory test: LDL Chol | 7 | 0 | **7** |
| South Korea KSR | Laboratory test: fasting glucose | 2 | 1 | **3** |
| Ontario OSR | Lab Investigations (subsequent tests to be considered following patient arrival to ED): HbA1C | 6 | 0 | **6** |
| South Korea KSR | Laboratory test: Cr | 6 | 1 | **7** |
| South Korea CRCS-5 | PT (Similar items aPTT & INR) | 4 | 0 | **4** |
| Scotland SSCA | Demographics: Ethnicity | 2 | 1 | **3** |

| | | | | |
|---|---|---|---|---|
| Scotland SSCA | Patient Pathway: Did the patient have a carotid intervention? | 1 | 2 | **3** |
| Scotland SSCA | Carotid Intervention: Date of carotid intervention | 2 | 1 | **3** |
| USA GWTG | First Glasgow Coma Scale (GCS) | 3 | 0 | **3** |
| USA PCNASP | Complications of thrombolytic therapy: Were there bleeding complications in a patient transferred after IV tPA | 1 | 2 | **3** |
| Austria ASUR | ASPECT score | 2 | 1 | **3** |

***Note.*** This table contains the items distilled from the 20 international audits/registries that do not appear in INAS, selected as they appear in 4 or more stroke audits/registries in total. Each row contains a reference item from the audit/registry of different countries and the item descriptor. In Phase 2 of the benchmarking exercise, each item was cross-checked against 19 other international audits/registries (excluding INAS) to see how frequently they ask the same [or closely related] similar item. These frequency counts are available for each item in the table. Note that these frequency counts do not include the index item itself within the count; hence, where the count says 3 that means 3 other audits/registries share the item in addition to the national audit from which the reference item is from. The items are colour-coded based on a traffic light system; whereby, dark green represents that the item is shared by a high number of registries and yellow marks that the item is shared with a moderate number of other registries.

Abbreviations: aPTT: activated partial thromboplastin clotting time; ASPECT: The Alberta Stroke Program Early CT Score; BP: blood pressure; Chol: cholesterol; Cr: creatinine; DBP: diastolic blood pressure; DVT: deep vein thrombosis; ECG: electrocardiogram; ED: emergency department; Hb: haemoglobin; HDL: high-density lipoprotein; IA: intra-arterial; ICH: intracerebral haemorrhage; INR: international normalised ratio; IV: intravenous; LPL: lipoprotein lipase; NIHSS: National Institutes of Health Stroke Scale; PT: prothrombin time; SAH: subarachnoid haemorrhage; SBP: systolic blood pressure; TG: triglycerides; TIA: transient ischaemic attack; TOAST: Trial of ORG 10172 in Acute Stroke Treatment; tPA: tissue plasminogen activator.

**Supplemental File F.** Results of the three-round Delphi Consultation regarding the retention of the INAS Core Clinical/Thrombectomy Items as part of the "Ideal" Core Minimum Acute Stroke Care Dataset for INAS

| AUDIT DETAILS | | DELPHI CONSULTATION | | | | | | | | | | | | | | Final Consensus |
|---|---|---|---|---|---|---|---|---|---|---|---|---|---|---|---|---|
| | | Round 1 Votes [a] | | | | | Round 2 Votes [b] | | | | | Round 3 Votes [c] | | | | |
| AUDIT | Item | % Include | % Exclude | % No Strong View | % Don't Know Enough | % Missing Data | % Include | % Exclude | % No Strong View | % Don't Know Enough | % Missing Data | % Include | % Exclude | % Don't Know Enough | % Missing Data | |
| **HIPE** | Age | **95** | 0 | 0 | 0 | 5 | **95** | 0 | 0 | 0 | 5 | | - | | | INCLUDE |
| **HIPE** | Date of discharge | **85** | 0 | 10 | 0 | 5 | **85** | 0 | 10 | 0 | 5 | | - | | | INCLUDE |
| **HIPE** | Hospital acquired diagnosis | **80** | 5 | 5 | 5 | 5 | **95** | 0 | 0 | 0 | 5 | | - | | | INCLUDE |
| **HIPE** | Length of stay | **90** | 0 | 5 | 0 | 5 | **95** | 0 | 0 | 0 | 5 | | - | | | INCLUDE |
| **HIPE** | Principal diagnosis | **95** | 0 | 0 | 0 | 5 | **95** | 0 | 0 | 0 | 5 | | - | | | INCLUDE |
| **HIPE** | Sex | **95** | 0 | 0 | 0 | 5 | **95** | 0 | 0 | 0 | 5 | | - | | | INCLUDE |
| **HIPE** | Source of admission | **85** | 5 | 5 | 0 | 5 | **90** | 0 | 5 | 0 | 5 | | - | | | INCLUDE |
| **HIPE** | Type of admission | 65 | 5 | 20 | 5 | 5 | **75** | 5 | 15 | 0 | 5 | | - | | | INCLUDE |
| **HIPE** | 1st additional diagnosis | **75** | 5 | 15 | 0 | 5 | **90** | 5 | 0 | 0 | 5 | | - | | | INCLUDE |
| **HIPE** | Hospital code list | **75** | 0 | 5 | 10 | 10 | **80** | 0 | 5 | 5 | 10 | | - | | | INCLUDE |
| **CORE CLINICAL** | Which hospital was patient transferred from (if any) | **80** | 0 | 5 | 5 | 10 | **85** | 0 | 5 | 0 | 10 | | - | | | INCLUDE |
| **CORE CLINICAL** | Why was the patient transferred? | **90** | 0 | 5 | 0 | 5 | **95** | 0 | 0 | 0 | 5 | | - | | | INCLUDE |
| **CORE CLINICAL** | If other transfer reason, please specify | 40 | 25 | 20 | 10 | 5 | 45 | 20 | 20 | 10 | 5 | | - | | | EXCLUDE |
| **CORE CLINICAL** | If other transfer hospital, please specify | 25 | 15 | 35 | 15 | 10 | 25 | 20 | 35 | 15 | 5 | | - | | | EXCLUDE |
| **CORE CLINICAL** | Symptom onset date | **95** | 0 | 0 | 0 | 5 | **95** | 0 | 0 | 0 | 5 | | - | | | INCLUDE |
| **CORE CLINICAL** | Symptom onset time | **95** | 0 | 0 | 0 | 5 | **95** | 0 | 0 | 0 | 5 | | - | | | INCLUDE |
| **CORE CLINICAL** | If symptom onset date is unknown, what date was the patient last known to be well | **75** | 10 | 5 | 5 | 5 | **80** | 5 | 5 | 5 | 5 | | - | | | INCLUDE |
| **CORE CLINICAL** | If symptom onset time is unknown, what time was the patient last known to be well | **75** | 10 | 5 | 5 | 5 | **80** | 5 | 5 | 5 | 5 | | - | | | INCLUDE |
| **CORE CLINICAL** | Did the stroke occur while the patient was in hospital for treatment of another condition | **85** | 5 | 0 | 5 | 5 | **90** | 0 | 0 | 5 | 5 | | - | | | INCLUDE |
| **CORE CLINICAL** | If no, date of presentation to hospital | **85** | 5 | 0 | 5 | 5 | **85** | 5 | 0 | 5 | 5 | | - | | | INCLUDE |

| | | | | | | | | | | | | | | | | |
|---|---|---|---|---|---|---|---|---|---|---|---|---|---|---|---|---|
| **CORE CLINICAL** | If no, time of presentation to hospital | **80** | 5 | 0 | 5 | 10 | **80** | 5 | 0 | 5 | 10 | | - | | | INCLUDE |
| **CORE CLINICAL** | If presentation time is unknown, was presentation to hospital within 4.5 hrs of symptom onset | 60 | 10 | 20 | 5 | 5 | 60 | 10 | 20 | 5 | 5 | 35 | 41 | 24 | 0 | EXCLUDE |
| **CORE CLINICAL** | Medical assessment date | 55 | 10 | 20 | 5 | 10 | 55 | 10 | 20 | 5 | 10 | 65 | 23 | 12 | 0 | EXCLUDE |
| **CORE CLINICAL** | Medical assessment time | 60 | 10 | 15 | 5 | 10 | 60 | 10 | 15 | 5 | 10 | 47 | 24 | 29 | 0 | EXCLUDE |
| **CORE CLINICAL** | Brain CT or MRI performed | **90** | 0 | 0 | 5 | 5 | **90** | 0 | 0 | 5 | 5 | | - | | | INCLUDE |
| **CORE CLINICAL** | If yes, First Brain Imaging date | **90** | 0 | 0 | 5 | 5 | **90** | 0 | 0 | 5 | 5 | | - | | | INCLUDE |
| **CORE CLINICAL** | If yes, First Brain Imaging time | **90** | 0 | 0 | 5 | 5 | **90** | 0 | 0 | 5 | 5 | | - | | | INCLUDE |
| **CORE CLINICAL** | Did the patient receive I.V. Thrombolysis | **90** | 0 | 0 | 5 | 5 | **90** | 0 | 0 | 5 | 5 | | - | | | INCLUDE |
| **CORE CLINICAL** | If yes, enter date | **90** | 0 | 0 | 5 | 5 | **90** | 0 | 0 | 5 | 5 | | - | | | INCLUDE |
| **CORE CLINICAL** | If yes, enter time | **90** | 0 | 0 | 5 | 5 | **90** | 0 | 0 | 5 | 5 | | - | | | INCLUDE |
| **CORE CLINICAL** | If yes, was intracerebral bleed seen on scan within 36 hrs | **80** | 0 | 5 | 10 | 5 | **85** | 0 | 5 | 5 | 5 | | - | | | INCLUDE |
| **CORE CLINICAL** | If intracerebral bleed, was neuro deterioration associated with it | **70** | 0 | 10 | 15 | 5 | **80** | 0 | 10 | 5 | 5 | | - | | | INCLUDE |
| **CORE CLINICAL** | Did the patient have thrombectomy in this hospital (Beaumont / CUH only) | **85** | 0 | 0 | 5 | 10 | **85** | 0 | 0 | 5 | 10 | | - | | | INCLUDE |
| **CORE CLINICAL** | Was a swallow screen completed | **90** | 0 | 0 | 5 | 5 | **90** | 0 | 0 | 5 | 5 | | - | | | INCLUDE |
| **CORE CLINICAL** | If yes, was swallow screen completed within 4 hours of presentation | **70** | 0 | 15 | 10 | 5 | **70** | 0 | 15 | 10 | 5 | | - | | | INCLUDE |
| **CORE CLINICAL** | Modified Rankin Scale pre-stroke | **85** | 0 | 5 | 5 | 5 | **85** | 0 | 5 | 5 | 5 | | - | | | INCLUDE |
| **CORE CLINICAL** | Admitted to Stroke Unit (Key Performance Indicator) | **90** | 0 | 0 | 5 | 5 | **90** | 0 | 0 | 5 | 5 | | - | | | INCLUDE |
| **CORE CLINICAL** | If yes, date admitted to Stroke Unit (Key Performance Indicator) | **90** | 0 | 0 | 5 | 5 | **90** | 0 | 0 | 5 | 5 | | - | | | INCLUDE |
| **CORE CLINICAL** | If yes, date discharged from Stroke Unit (Key Performance Indicator) | **80** | 5 | 0 | 5 | 10 | **85** | 0 | 0 | 5 | 10 | | - | | | INCLUDE |
| **CORE CLINICAL** | If no, reason why | **75** | 0 | 10 | 5 | 10 | **80** | 0 | 5 | 5 | 10 | | - | | | INCLUDE |
| **CORE CLINICAL** | If other reason, please specify | 55 | 15 | 15 | 5 | 10 | 55 | 15 | 15 | 5 | 10 | 58 | 18 | 18 | 6 | EXCLUDE |
| **CORE CLINICAL** | Allied Health Professional (AHP) Assessment | **80** | 5 | 5 | 5 | 5 | **85** | 5 | 0 | 5 | 5 | | - | | | INCLUDE |
| **CORE CLINICAL** | If yes, Physiotherapy | **80** | 5 | 5 | 5 | 5 | **85** | 5 | 0 | 5 | 5 | | - | | | INCLUDE |
| **CORE CLINICAL** | If yes, Occupational Therapy | **80** | 5 | 5 | 5 | 5 | **85** | 5 | 0 | 5 | 5 | | - | | | INCLUDE |
| **CORE CLINICAL** | If yes, Speech and Language Therapy | **80** | 5 | 5 | 5 | 5 | **85** | 5 | 0 | 5 | 5 | | - | | | INCLUDE |
| **CORE CLINICAL** | If yes, Dietetics | **75** | 5 | 5 | 10 | 5 | **80** | 5 | 0 | 10 | 5 | | - | | | INCLUDE |

| | | | | | | | | | | | | | | | |
|---|---|---|---|---|---|---|---|---|---|---|---|---|---|---|---|
| **CORE CLINICAL** | If yes, Medical Social Worker | **75** | 5 | 5 | 10 | 5 | **80** | 5 | 0 | 10 | 5 | | - | | | INCLUDE |
| **CORE CLINICAL** | If yes, Psychology | **75** | 5 | 5 | 10 | 5 | **80** | 5 | 0 | 10 | 5 | | - | | | INCLUDE |
| **CORE CLINICAL** | Was the patient assessed by Stroke Nurse Specialist | **75** | 0 | 10 | 10 | 5 | **80** | 0 | 5 | 10 | 5 | | - | | | INCLUDE |
| **CORE CLINICAL** | If no, reason why | 45 | 15 | 20 | 10 | 10 | 50 | 15 | 15 | 10 | 10 | 65 | 23 | 6 | 6 | EXCLUDE |
| **CORE CLINICAL** | Multidisciplinary Meeting Case assessment | 50 | 10 | 20 | 15 | 5 | 55 | 10 | 15 | 15 | 5 | 41 | 35 | 24 | 0 | EXCLUDE |
| **CORE CLINICAL** | Was an assessment of mood completed and documented by a member of the multidisciplinary team | **75** | 0 | 10 | 10 | 5 | **80** | 0 | 5 | 10 | 5 | | - | | | INCLUDE |
| **CORE CLINICAL** | Does the patient have Symptomatic Carotid Stenosis | 50 | 0 | 15 | 30 | 5 | 50 | 0 | 15 | 30 | 5 | 65 | 0 | 35 | 0 | EXCLUDE |
| **CORE CLINICAL** | If Symptomatic Carotid Stenosis, was the patient referred for Carotid Endarterectomy | 50 | 5 | 10 | 30 | 5 | 50 | 5 | 10 | 30 | 5 | 59 | 6 | 35 | 0 | EXCLUDE |
| **CORE CLINICAL** | If Symptomatic Carotid Stenosis, was the patient referred for Carotid Stenting | 50 | 5 | 10 | 30 | 5 | 50 | 5 | 10 | 30 | 5 | 53 | 6 | 41 | 0 | EXCLUDE |
| **CORE CLINICAL** | New or Altered Antithrombotic Therapy prescribed for acute treatment | 65 | 0 | 10 | 20 | 5 | **70** | 0 | 10 | 15 | 5 | | - | | | INCLUDE |
| **CORE CLINICAL** | If yes, Antiplatelet or Anticoagulant (for acute treatment) start date | 60 | 5 | 5 | 20 | 10 | **70** | 5 | 5 | 15 | 5 | | - | | | INCLUDE |
| **CORE CLINICAL** | Does the patient have Atrial Fibrillation | **80** | 0 | 0 | 15 | 5 | **80** | 0 | 0 | 15 | 5 | | - | | | INCLUDE |
| **CORE CLINICAL** | If Atrial Fibrillation, was atrial fibrillation known prior to stroke onset | **80** | 0 | 0 | 15 | 5 | **80** | 0 | 0 | 15 | 5 | | - | | | INCLUDE |
| **CORE CLINICAL** | If atrial fibrillation known prior to stroke onset, was Antiplatelet and/or Anticoagulant prescribed prior to Stroke onset | **70** | 5 | 0 | 20 | 5 | **75** | 5 | 0 | 15 | 5 | | - | | | INCLUDE |
| **CORE CLINICAL** | If yes, please specify Antiplatelet / Anticoagulant - prior to stroke | **75** | 0 | 0 | 20 | 5 | **80** | 0 | 0 | 15 | 5 | | - | | | INCLUDE |
| **CORE CLINICAL** | If atrial fibrillation known prior to stroke onset, and on Warfarin, was INR (International Normalised Ratio) 2-3 at Stroke onset. | 60 | 5 | 5 | 20 | 10 | 65 | 5 | 5 | 20 | 5 | **70** | 12 | 18 | 0 | INCLUDE |
| **CORE CLINICAL** | If Atrial Fibrillation, Anticoagulation prescribed for secondary prevention | **75** | 0 | 0 | 15 | 10 | **80** | 0 | 0 | 15 | 5 | | - | | | INCLUDE |
| **CORE CLINICAL** | If yes, please specify Antiplatelet / Anticoagulant - on discharge | **70** | 0 | 5 | 10 | 10 | **75** | 0 | 5 | 15 | 5 | | - | | | INCLUDE |
| **CORE CLINICAL** | If no, please enter reason documented | 60 | 0 | 10 | 20 | 10 | 65 | 0 | 10 | 20 | 5 | **70** | 12 | 12 | 6 | INCLUDE |
| **CORE CLINICAL** | Modified Rankin Scale on discharge | **75** | 0 | 10 | 5 | 10 | **80** | 0 | 10 | 5 | 5 | | - | | | INCLUDE |
| **CORE CLINICAL** | Discharge destination | **90** | 0 | 0 | 0 | 10 | **95** | 0 | 0 | 0 | 5 | | - | | | INCLUDE |
| **CORE CLINICAL** | Case complete | 30 | 20 | 25 | 15 | 10 | 30 | 20 | 25 | 20 | 5 | | - | | | EXCLUDE |

| | | | | | | | | | | | | | | | | |
|---|---|---|---|---|---|---|---|---|---|---|---|---|---|---|---|---|
| **THROMBECTOMY** | NIHSS pre-thrombectomy | **80** | 0 | 5 | 5 | 10 | **85** | 0 | 5 | 5 | 5 | - | | | | INCLUDE |
| **THROMBECTOMY** | Date of performance of non-contrast CT | 55 | 5 | 10 | 20 | 10 | 55 | 5 | 10 | 25 | 5 | 47 | 12 | 35 | 6 | EXCLUDE |
| **THROMBECTOMY** | Time of performance of non-contrast CT | 55 | 5 | 10 | 20 | 10 | 55 | 5 | 10 | 25 | 5 | 47 | 6 | 41 | 6 | EXCLUDE |
| **THROMBECTOMY** | Date of performance of non-contrast CTA | 60 | 0 | 10 | 20 | 10 | 60 | 0 | 10 | 25 | 5 | 41 | 12 | 41 | 6 | EXCLUDE |
| **THROMBECTOMY** | Time of performance of non-contrast CTA | 60 | 0 | 10 | 20 | 10 | 60 | 0 | 10 | 25 | 5 | 41 | 12 | 41 | 6 | EXCLUDE |
| **THROMBECTOMY** | Date of contact with the endovascular stroke centre | 60 | 5 | 5 | 20 | 10 | 60 | 5 | 5 | 25 | 5 | 47 | 6 | 41 | 6 | EXCLUDE |
| **THROMBECTOMY** | Time of contact with the endovascular stroke centre | 60 | 5 | 5 | 20 | 10 | 60 | 5 | 5 | 25 | 5 | 47 | 6 | 41 | 6 | EXCLUDE |
| **THROMBECTOMY** | Date of decision to transfer patient | **70** | 0 | 5 | 15 | 10 | **75** | 0 | 5 | 15 | 5 | - | | | | INCLUDE |
| **THROMBECTOMY** | Time of decision to transfer patient | 60 | 0 | 15 | 15 | 10 | 65 | 0 | 15 | 15 | 5 | 59 | 0 | 35 | 6 | EXCLUDE |
| **THROMBECTOMY** | Was the patient transferred from another hospital | 60 | 0 | 15 | 15 | 10 | **70** | 0 | 10 | 15 | 5 | - | | | | INCLUDE |
| **THROMBECTOMY** | If yes, what date did the patient arrive at the referring/first hospital | 65 | 0 | 10 | 15 | 10 | **70** | 0 | 10 | 15 | 5 | - | | | | INCLUDE |
| **THROMBECTOMY** | If yes, what time did the patient arrive at the referring/first hospital | 65 | 0 | 10 | 15 | 10 | 65 | 0 | 10 | 20 | 5 | 47 | 6 | 41 | 6 | EXCLUDE |
| **THROMBECTOMY** | If yes, what date did the patient leave the referring/first hospital for transfer to the EVT centre? | 55 | 0 | 20 | 15 | 10 | 55 | 0 | 20 | 20 | 5 | 47 | 6 | 41 | 6 | EXCLUDE |
| **THROMBECTOMY** | If yes, what time did the patient leave the referring/first hospital for transfer to the EVT centre? | 55 | 0 | 20 | 15 | 10 | 55 | 0 | 20 | 20 | 5 | 53 | 6 | 35 | 6 | EXCLUDE |
| **THROMBECTOMY** | Date of arrival at the endovascular stroke centre | 60 | 0 | 15 | 15 | 10 | 60 | 0 | 15 | 20 | 5 | 59 | 0 | 29 | 12 | EXCLUDE |
| **THROMBECTOMY** | Time of arrival at the endovascular stroke centre | 60 | 0 | 15 | 15 | 10 | 60 | 0 | 15 | 20 | 5 | 53 | 0 | 41 | 6 | EXCLUDE |
| **THROMBECTOMY** | Did the patient have repeat non-invasive imaging in the endovascular stroke centre | 50 | 5 | 20 | 15 | 10 | 50 | 10 | 15 | 20 | 5 | 41 | 18 | 35 | 6 | EXCLUDE |
| **THROMBECTOMY** | If yes, please specify | 45 | 5 | 25 | 15 | 10 | 45 | 10 | 20 | 20 | 5 | 41 | 18 | 35 | 6 | EXCLUDE |
| **THROMBECTOMY** | Site of most proximal occlusion | **70** | 0 | 10 | 10 | 10 | **75** | 5 | 5 | 10 | 5 | - | | | | INCLUDE |
| **THROMBECTOMY** | Second occlusion site | 45 | 5 | 25 | 15 | 10 | 45 | 10 | 20 | 20 | 5 | 35 | 24 | 35 | 6 | EXCLUDE |
| **THROMBECTOMY** | Associated carotid stenosis greater than 50% | 60 | 0 | 15 | 15 | 10 | 65 | 5 | 10 | 15 | 5 | 41 | 18 | 35 | 6 | EXCLUDE |
| **THROMBECTOMY** | TICI pre thrombectomy | 55 | 0 | 10 | 25 | 10 | 50 | 5 | 10 | 30 | 5 | 47 | 12 | 35 | 6 | EXCLUDE |
| **THROMBECTOMY** | TICI post thrombectomy | 60 | 0 | 10 | 20 | 10 | 55 | 5 | 10 | 25 | 5 | 47 | 12 | 35 | 6 | EXCLUDE |
| **THROMBECTOMY** | Date of groin puncture | 50 | 0 | 15 | 25 | 10 | 50 | 5 | 10 | 30 | 5 | 41 | 18 | 35 | 6 | EXCLUDE |
| **THROMBECTOMY** | Time of groin puncture | 50 | 0 | 15 | 25 | 10 | 50 | 5 | 10 | 30 | 5 | 41 | 18 | 35 | 6 | EXCLUDE |
| **THROMBECTOMY** | Date of first pass | 30 | 5 | 15 | 40 | 10 | 35 | 10 | 5 | 45 | 5 | 35 | 24 | 35 | 6 | EXCLUDE |

| | | | | | | | | | | | | | | | |
|---|---|---|---|---|---|---|---|---|---|---|---|---|---|---|---|
| **THROMBECTOMY** | Time of first pass | 30 | 5 | 15 | 40 | 10 | 35 | 10 | 5 | 45 | 5 | 35 | 24 | 35 | 6 | EXCLUDE |
| **THROMBECTOMY** | Date of first reperfusion | 40 | 5 | 15 | 30 | 10 | 40 | 10 | 10 | 35 | 5 | 41 | 18 | 35 | 6 | EXCLUDE |
| **THROMBECTOMY** | Time of first reperfusion | 40 | 5 | 15 | 30 | 10 | 40 | 10 | 10 | 35 | 5 | 41 | 18 | 35 | 6 | EXCLUDE |
| **THROMBECTOMY** | Date of final angio | 35 | 5 | 15 | 35 | 10 | 35 | 10 | 10 | 40 | 5 | 29 | 29 | 35 | 6 | EXCLUDE |
| **THROMBECTOMY** | Time of final angio | 35 | 5 | 15 | 35 | 10 | 35 | 10 | 10 | 40 | 5 | 29.5 | 29.5 | 35 | 6 | EXCLUDE |
| **THROMBECTOMY** | Immediate complications | **70** | 0 | 10 | 10 | 10 | **70** | 0 | 10 | 15 | 5 | - | | | | INCLUDE |
| **THROMBECTOMY** | NIHSS 24-hour post-thrombectomy | **75** | 0 | 5 | 10 | 10 | **75** | 0 | 5 | 15 | 5 | - | | | | INCLUDE |
| **THROMBECTOMY** | Following procedure was patient transferred immediately back to primary receiving hospital | 65 | 0 | 10 | 15 | 10 | 65 | 0 | 10 | 20 | 5 | 65 | 0 | 29 | 6 | EXCLUDE |
| **THROMBECTOMY** | If no, when was patient admitted to the endovascular stroke centre | 55 | 5 | 10 | 20 | 10 | 50 | 5 | 15 | 25 | 5 | 59 | 6 | 29 | 6 | EXCLUDE |

Note. The emboldened numbers represent those items that received ≥70% of the votes to Include as part of INAS. The numbers in red font represent those items for which there were vote changes made from Round 1 to Round 2. A reduced set of items were put forward for review in Round 3.

[a] Twenty out of 24 eligible stakeholders responded with item votes in the first round of the Delphi Consultation, representing an 83% response rate. The percentages represent the percentage of the 20 responding stakeholders who voted for each response option.

[b] Eleven out of the initially responding 20 stakeholders (55% response rate) responded with changed responses during the second round of the Delphi consultation. On average, 4.91% of the responses were changed by responding stakeholders; that is, they changed their vote from one eligible response to another. An additional 6.55% of responses were changed from an originally 'missing' vote to one of the eligible response options. Eight stakeholders did not respond, interpreted as no change to their previous responses. One additional stakeholder left their post after Round 1 and so their previous responses remained unchanged. The percentages displayed in the table represent the combined percentage votes across Rounds 1 and 2 for the 20 responding stakeholders including the initial votes for the 9 stakeholders who made no changes and the amended votes for the 11 responding participants in Round 2.
[c] The percentages represent the Delphi Round 3 votes for a reduced set of items and response options from 17 responding participants out of 22 eligible stakeholders (77% response rate).

Abbreviations: Angio: angiogram; CT: computed tomography; CTA: computed tomography angiography; CUH: Cork University Hospital; EVT: endovascular thrombectomy; HIPE: Hospital In-Patient Enquiry; IV: intravenous; MRI: magnetic resonance imaging; NIHSS: National Institutes of Health Stroke Scale; TICI: thrombolysis in cerebral infarction.

**Supplemental File G.** Results of the three-round Delphi Consultation regarding the recommended inclusion of frequently collected international Items and additional expert-suggested items as part of the "Ideal" Core Minimum Acute Stroke Care Dataset for INAS

| AUDIT DETAILS | | ROUND 1 VOTES [A] | | | | | ROUND 2 VOTES [B] | | | | | ROUND 3 VOTES [C] | | | | |
|---|---|---|---|---|---|---|---|---|---|---|---|---|---|---|---|---|
| **COUNTRY OF AUDIT** | **International Audit Reference Item Descriptor** | **% Include** | **% Exclude** | **% No Strong View** | **% Don't Know Enough** | **% Missing Data** | **% Include** | **% Exclude** | **% No Strong View** | **% Don't Know Enough** | **% Missing Data** | **% Include** | **% Exclude** | **% Don't Know Enough** | **% Missing Data** | **Final Consensus** |
| **AUSTRALIA AUSCR** | Patient Record ID [Patient Details] | 50 | 25 | 10 | 0 | 15 | 60 | 25 | 10 | 0 | 5 | **70** | 18 | 12 | 0 | INCLUDE |
| **AUSTRALIA AUSCR** | First name [Patient Details] | 35 | 40 | 10 | 5 | 10 | 45 | 40 | 10 | 5 | 0 | - | | | | EXCLUDE |
| **AUSTRALIA AUSCR** | Last name [Patient Details] | 35 | 45 | 5 | 5 | 10 | 45 | 45 | 5 | 5 | 0 | - | | | | EXCLUDE |
| **AUSTRALIA AUSCR** | Country of birth [Patient Details] | 45 | 40 | 10 | 0 | 5 | 45 | 40 | 15 | 0 | 0 | - | | | | EXCLUDE |
| **AUSTRALIA AUSCR** | Postcode [Contact Information] | 50 | 45 | 0 | 0 | 5 | 55 | 45 | 0 | 0 | 0 | 41 | 41 | 18 | 0 | EXCLUDE |
| **AUSTRALIA AUSCR** | Transport code [Pre-Hospital Care] | 25 | 35 | 5 | 30 | 5 | 25 | 40 | 10 | 25 | 0 | - | | | | EXCLUDE |
| **AUSTRALIA AUSCR** | Did the patient arrive by ambulance? [Admission Details] | **70** | 20 | 0 | 5 | 5 | **75** | 20 | 0 | 5 | 0 | **82** | 12 | 6 | 0 | INCLUDE |
| **AUSTRALIA AUSCR** | Previous stroke (A history of stroke prior to this current episode, excluding TIAs) [History of Known Risk Factors] | **80** | 10 | 0 | 5 | 5 | **85** | 10 | 0 | 5 | 0 | **88** | 12 | 0 | 0 | INCLUDE |
| **AUSTRALIA AUSCR** | Previous TIA (Transient Ischemic Attack (TIA) or mini stroke) [History of Known Risk Factors] | **70** | 20 | 0 | 5 | 5 | **75** | 20 | 0 | 5 | 0 | **88** | 12 | 0 | 0 | INCLUDE |
| **AUSTRALIA AUSCR** | Diabetes [History of Known Risk Factors] | **90** | 0 | 0 | 5 | 5 | **95** | 0 | 0 | 5 | 0 | **88** | 12 | 0 | 0 | INCLUDE |
| **AUSTRALIA AUSCR** | Hypercholesterolaemia [History of Known Risk Factors] | **90** | 0 | 0 | 5 | 5 | **95** | 0 | 0 | 5 | 0 | **88** | 12 | 0 | 0 | INCLUDE |
| **AUSTRALIA AUSCR** | Hypertension [History of Known Risk Factors] | **90** | 0 | 0 | 5 | 5 | **95** | 0 | 0 | 5 | 0 | **88** | 12 | 0 | 0 | INCLUDE |
| **AUSTRALIA AUSCR** | Recent myocardial infarction [History of Known Risk Factors] | 65 | 15 | 5 | 10 | 5 | **70** | 15 | 5 | 10 | 0 | **76** | 12 | 12 | 0 | INCLUDE |
| **AUSTRALIA AUSCR** | Ischaemic heart disease [History of Known Risk Factors] | **90** | 0 | 0 | 5 | 5 | **95** | 0 | 0 | 5 | 0 | **82** | 12 | 6 | 0 | INCLUDE |
| **AUSTRALIA AUSCR** | High alcohol consumption [History of Known Risk Factors] | **85** | 5 | 5 | 0 | 5 | **90** | 5 | 5 | 0 | 0 | **76** | 18 | 6 | 0 | INCLUDE |

| | | | | | | | | | | | | | | | | |
|---|---|---|---|---|---|---|---|---|---|---|---|---|---|---|---|---|
| **AUSTRALIA AUSCR** | Current smoker [History of Known Risk Factors] | **95** | 0 | 0 | 0 | 5 | **100** | 0 | 0 | 0 | 0 | **88** | 12 | 0 | 0 | INCLUDE |
| **AUSTRALIA AUSCR** | Past smoker [History of Known Risk Factors] | **80** | 10 | 5 | 0 | 5 | **90** | 5 | 5 | 0 | 0 | **71** | 23 | 6 | 0 | INCLUDE |
| **AUSTRALIA AUSCR** | Family history of stroke [History of Known Risk Factors] | 65 | 10 | 15 | 5 | 5 | **75** | 10 | 10 | 5 | 0 | 59 | 29 | 12 | 0 | EXCLUDE |
| **AUSTRALIA AUSCR** | Obesity [History of Known Risk Factors] | **70** | 5 | 15 | 0 | 10 | **80** | 5 | 10 | 0 | 5 | **82** | 18 | 0 | 0 | INCLUDE |
| **AUSTRALIA AUSCR** | Peripheral vascular disease [History of Known Risk Factors] | **75** | 10 | 5 | 5 | 5 | **80** | 10 | 5 | 5 | 0 | **70** | 18 | 12 | 0 | INCLUDE |
| **AUSTRALIA AUSCR** | Other serious illness that influences prognosis or management of stroke [History of Known Risk Factors] | 60 | 10 | 25 | 0 | 5 | 65 | 10 | 25 | 0 | 0 | 59 | 23 | 18 | 0 | EXCLUDE |
| **AUSTRALIA AUSCR** | Did the patient take antihypertensives prior to this stroke? [Pre-Admission Medication] | 55 | 5 | 20 | 15 | 5 | 60 | 5 | 20 | 15 | 0 | **76** | 24 | 0 | 0 | INCLUDE |
| **AUSTRALIA AUSCR** | Did the patient take lipid lowering treatment prior to this stroke? [Pre-Admission Medication] | 50 | 5 | 20 | 20 | 5 | 55 | 5 | 20 | 20 | 0 | **71** | 23 | 6 | 0 | INCLUDE |
| **AUSTRALIA AUSCR** | Statin [Pre-Admission Medication] | 55 | 5 | 25 | 10 | 5 | 60 | 5 | 25 | 10 | 0 | 59 | 35 | 6 | 0 | EXCLUDE |
| **AUSTRALIA AUSCR** | Can the patient walk on their own (i.e. without the assistance of another person, but may include walking aid)? - Pre-stroke functional status. [Dependency Prior to Admission] | 45 | 25 | 20 | 5 | 5 | 55 | 25 | 15 | 5 | 0 | 59 | 23 | 18 | 0 | EXCLUDE |
| **AUSTRALIA AUSCR** | Living arrangements prior to admission [Dependency Prior to Admission] | 45 | 25 | 25 | 0 | 5 | 55 | 25 | 20 | 0 | 0 | 47 | 47 | 6 | 0 | EXCLUDE |
| **AUSTRALIA AUSCR** | Assessment for level of responsiveness [Acute Clinical Data] | 35 | 40 | 10 | 10 | 5 | 40 | 40 | 10 | 10 | 0 | - | | | | EXCLUDE |
| **AUSTRALIA AUSCR** | Was the patient unresponsive (or in a coma)? [Acute Clinical Data] | 25 | 45 | 10 | 15 | 5 | 30 | 45 | 10 | 15 | 0 | - | | | | EXCLUDE |
| **AUSTRALIA AUSCR** | Patient's glucose level before thrombolysis treatment or on hospital arrival (if no thrombolysis) amount (mmol/l) [Acute Clinical Data] | 35 | 20 | 10 | 30 | 5 | 35 | 20 | 15 | 30 | 0 | - | | | | EXCLUDE |
| **AUSTRALIA AUSCR** | NIHSS at baseline [Acute Clinical Data] | **70** | 15 | 0 | 10 | 5 | **75** | 15 | 0 | 10 | 0 | **76** | 6 | 18 | 0 | INCLUDE |
| **AUSTRALIA AUSCR** | Weight at baseline, Amount (kg) [Acute Clinical Data] | 55 | 25 | 5 | 10 | 5 | 60 | 25 | 5 | 10 | 0 | 53 | 41 | 6 | 0 | EXCLUDE |
| **AUSTRALIA AUSCR** | BP at baseline (SBP/DBP) [Acute Clinical Data] | **70** | 20 | 0 | 5 | 5 | **75** | 20 | 0 | 5 | 0 | 65 | 29 | 6 | 0 | EXCLUDE |

| | | | | | | | | | | | | | | | | |
|---|---|---|---|---|---|---|---|---|---|---|---|---|---|---|---|---|
| **AUSTRALIA AUSCR** | What type of brain scan was performed? [Brain Imaging] | 40 | 20 | 15 | 20 | 5 | 45 | 20 | 15 | 20 | 0 | - | | | | EXCLUDE |
| **AUSTRALIA AUSCR** | Type of stroke [Brain Imaging] | 50 | 20 | 5 | 20 | 5 | 55 | 20 | 5 | 20 | 0 | 47 | 35 | 18 | 0 | EXCLUDE |
| **AUSTRALIA AUSCR** | Side of stroke affected [Brain Imaging] | 50 | 10 | 20 | 15 | 5 | 55 | 10 | 20 | 15 | 0 | 41 | 35 | 24 | 0 | EXCLUDE |
| **AUSTRALIA AUSCR** | Cause of stroke (Stroke cause determined based on TOAST classification system) [Brain Imaging] | 55 | 15 | 5 | 20 | 5 | 60 | 15 | 5 | 20 | 0 | 59 | 29 | 12 | 0 | EXCLUDE |
| **AUSTRALIA AUSCR** | Mechanism (The cause or mechanism of current stroke episode.) [Brain Imaging] | 35 | 25 | 10 | 25 | 5 | 35 | 25 | 10 | 30 | 0 | - | | | | EXCLUDE |
| **AUSTRALIA AUSCR** | Did the patient have a 12 lead ECG in hospital? [Other Investigations] | 25 | 30 | 10 | 25 | 10 | 25 | 30 | 10 | 30 | 5 | - | | | | EXCLUDE |
| **AUSTRALIA AUSCR** | Type of other cardiac investigations - If yes, specify [Other Investigations] | 30 | 20 | 20 | 25 | 5 | 35 | 20 | 20 | 25 | 0 | - | | | | EXCLUDE |
| **AUSTRALIA AUSCR** | Drug (Drug used for intravenous thrombolysis) [Telemedicine Setting and Reason] | 35 | 20 | 0 | 40 | 5 | 35 | 20 | 5 | 40 | 0 | - | | | | EXCLUDE |
| **AUSTRALIA AUSCR** | Dose (Full (0.9mg/kg)) (The total number of milligrams of intravenous thrombolysis therapy given intravenously (combined amount of bolus and infusion amount)) [Telemedicine Setting and Reason] | 20 | 30 | 5 | 40 | 5 | 25 | 30 | 5 | 40 | 0 | - | | | | EXCLUDE |
| **AUSTRALIA AUSCR** | Was patient outside of time window? (Patient was outside of the window of time when thrombolysis can safely be given) [Telemedicine Setting and Reason] | 40 | 20 | 5 | 30 | 5 | 40 | 25 | 5 | 30 | 0 | - | | | | EXCLUDE |
| **AUSTRALIA AUSCR** | Patient was fully worked up within 4.5 hours but tPA unable to be given for following reasons [Telemedicine Setting and Reason] | 35 | 20 | 15 | 25 | 5 | 30 | 30 | 15 | 25 | 0 | - | | | | EXCLUDE |
| **AUSTRALIA AUSCR** | Age> 80 (Reason(s) why thrombolysis was not given (despite patient being fully worked up within 4.5 hours).) [Telemedicine Setting and Reason] | 25 | 35 | 10 | 25 | 5 | 30 | 35 | 10 | 25 | 0 | - | | | | EXCLUDE |
| **AUSTRALIA AUSCR** | Major co-morbidity (Reason(s) why thrombolysis was not given (despite patient being fully worked up within 4.5 hours).) [Telemedicine Setting and Reason] | 30 | 25 | 15 | 25 | 5 | 35 | 25 | 15 | 25 | 0 | - | | | | EXCLUDE |
| **AUSTRALIA AUSCR** | Warfarin with INR> 1.7 (Reason(s) why thrombolysis was not given (despite patient being fully worked | 25 | 35 | 5 | 30 | 5 | 30 | 35 | 5 | 30 | 0 | - | | | | EXCLUDE |

| | | | | | | | | | | | | | | | | |
|---|---|---|---|---|---|---|---|---|---|---|---|---|---|---|---|---|
| | up within 4.5 hours).) [Telemedicine Setting and Reason] | | | | | | | | | | | | | | | |
| **AUSTRALIA AUSCR** | Rapidly improving (Reason(s) why thrombolysis was not given (despite patient being fully worked up within 4.5 hours).) [Telemedicine Setting and Reason] | 30 | 30 | 5 | 30 | 5 | 35 | 30 | 5 | 30 | 0 | - | | | | EXCLUDE |
| **AUSTRALIA AUSCR** | Other contraindication (Reason(s) why thrombolysis was not given (despite patient being fully worked up within 4.5 hours).) [Telemedicine Setting and Reason] | 30 | 25 | 10 | 30 | 5 | 35 | 25 | 10 | 30 | 0 | - | | | | EXCLUDE |
| **AUSTRALIA AUSCR** | Anaesthesia used (Type of anaesthesia used during reperfusion treatment) [Telemedicine Setting and Reason] | 20 | 25 | 15 | 35 | 5 | 20 | 25 | 20 | 35 | 0 | - | | | | EXCLUDE |
| **AUSTRALIA AUSCR** | Primary/Initial IA treatment (Primary thrombectomy method/device used) [Telemedicine Setting and Reason] | 15 | 30 | 15 | 35 | 5 | 15 | 30 | 20 | 35 | 0 | - | | | | EXCLUDE |
| **AUSTRALIA AUSCR** | Heparin (Heparin used during endovascular treatment.) [Telemedicine Setting and Reason] | 10 | 35 | 15 | 35 | 5 | 10 | 35 | 20 | 35 | 0 | - | | | | EXCLUDE |
| **AUSTRALIA AUSCR** | If yes, were fluids administered via IV or nasogastric tube? [Hydration and Nutrition] | 5 | 40 | 10 | 40 | 5 | 5 | 40 | 15 | 40 | 0 | - | | | | EXCLUDE |
| **AUSTRALIA AUSCR** | Did the patient with intracerebral haemorrhage receive hemicraniectomy [Surgical Management] | 55 | 10 | 0 | 30 | 5 | 60 | 10 | 0 | 30 | 0 | 53 | 23.5 | 23.5 | 0 | EXCLUDE |
| **AUSTRALIA AUSCR** | Date of hemicraniectomy [Surgical Management] | 40 | 10 | 15 | 30 | 5 | 40 | 15 | 10 | 35 | 0 | - | | | | EXCLUDE |
| **AUSTRALIA AUSCR** | Was a venous blood glucose level sample collected and sent to laboratory while patient was in the ED? [Assessment and Management of Hyperglycaemia] | 10 | 45 | 0 | 40 | 5 | 10 | 45 | 0 | 40 | 5 | - | | | | EXCLUDE |
| **AUSTRALIA AUSCR** | Did management of the patient include Heparin (or Low Molecular Weight Heparin)? [DVT Prophylaxis] | 15 | 35 | 5 | 40 | 5 | 15 | 35 | 10 | 40 | 0 | - | | | | EXCLUDE |
| **AUSTRALIA AUSCR** | Aspiration pneumonia [Complications during Hospital Admission] | 60 | 10 | 0 | 25 | 5 | 65 | 10 | 0 | 25 | 0 | **94** | 6 | 0 | 0 | INCLUDE |
| **AUSTRALIA AUSCR** | Deep Vein Thrombosis (DVT) [Complications during Hospital Admission] | 60 | 5 | 5 | 25 | 5 | 65 | 5 | 5 | 25 | 0 | **94** | 6 | 0 | 0 | INCLUDE |

| | | | | | | | | | | | | | | | | |
|---|---|---|---|---|---|---|---|---|---|---|---|---|---|---|---|---|
| **AUSTRALIA AUSCR** | Pulmonary Thrombo-Embolism (PE) [Complications during Hospital Admission] | 65 | 5 | 5 | 20 | 5 | **70** | 5 | 5 | 20 | 0 | **70** | 12 | 18 | 0 | INCLUDE |
| **AUSTRALIA AUSCR** | Acute myocardial infarction [Complications during Hospital Admission] | 65 | 10 | 0 | 20 | 5 | **70** | 10 | 0 | 20 | 0 | 65 | 17.5 | 17.5 | 0 | EXCLUDE |
| **AUSTRALIA AUSCR** | New stroke [Complications during Hospital Admission] | **70** | 5 | 5 | 15 | 5 | **80** | 0 | 5 | 15 | 0 | **82** | 12 | 6 | 0 | INCLUDE |
| **AUSTRALIA AUSCR** | Urinary tract infection [Complications during Hospital Admission] | **70** | 10 | 0 | 15 | 5 | **75** | 10 | 0 | 15 | 0 | **82** | 12 | 6 | 0 | INCLUDE |
| **AUSTRALIA AUSCR** | Seizures [Complications during Hospital Admission] | 65 | 5 | 5 | 20 | 5 | **75** | 0 | 5 | 20 | 0 | **82** | 18 | 0 | 0 | INCLUDE |
| **AUSTRALIA AUSCR** | Other [Complications during Hospital Admission] | 55 | 15 | 0 | 25 | 5 | 60 | 15 | 0 | 25 | 0 | 41 | 29 | 18 | 12 | EXCLUDE |
| **AUSTRALIA AUSCR** | On discharge was the patient prescribed antihypertensive agents? [Medication Prescribed at Discharge] | 60 | 5 | 5 | 25 | 5 | 65 | 5 | 5 | 25 | 0 | **70** | 18 | 12 | 0 | INCLUDE |
| **AUSTRALIA AUSCR** | On discharge was the patient prescribed lipid lowering treatment? [Medication Prescribed at Discharge] | 60 | 5 | 5 | 25 | 5 | 65 | 5 | 5 | 25 | 0 | 65 | 17.5 | 17.5 | 0 | EXCLUDE |
| **AUSTRALIA AUSCR** | Statin [Medication Prescribed at Discharge] | 45 | 25 | 10 | 15 | 5 | 45 | 25 | 15 | 15 | 0 | - | | | | EXCLUDE |
| **AUSTRALIA AUSCR** | Patient deceased during hospital care? [Discharge Information] | **75** | 5 | 10 | 5 | 5 | **80** | 5 | 10 | 5 | 0 | 59 | 41 | 0 | 0 | EXCLUDE |
| **AUSTRALIA AUSCR** | Date of death [Discharge Information] | 65 | 20 | 5 | 5 | 5 | **70** | 20 | 5 | 5 | 0 | 59 | 35 | 0 | 6 | EXCLUDE |
| **AUSTRALIA AUSCR** | Cause of death [Discharge Information] | 65 | 15 | 5 | 10 | 5 | **70** | 15 | 5 | 10 | 0 | 53 | 35 | 12 | 0 | EXCLUDE |
| **AUSTRALIA AUSCR** | NIHSS at discharge [Dependency on Discharge] | 55 | 20 | 5 | 15 | 5 | 60 | 20 | 5 | 15 | 0 | 59 | 29 | 12 | 0 | EXCLUDE |
| **AUSTRALIA AUSCR** | Date of NIHSS if not at discharge (e.g. day 2-7) [Dependency on Discharge] | 40 | 15 | 25 | 15 | 5 | 40 | 20 | 25 | 15 | 0 | - | | | | EXCLUDE |
| **AUSTRALIA AUSCR** | Death date post-acute episode of care [Record Death for Patient] | 45 | 20 | 15 | 15 | 5 | 50 | 20 | 15 | 15 | 0 | 65 | 35 | 0 | 0 | EXCLUDE |
| **SWEDEN RIKSSTROKE** | Enter hospital code where thrombolysis was performed [Thrombolysis] | 15 | 50 | 5 | 25 | 5 | 15 | 50 | 5 | 30 | 0 | - | | | | EXCLUDE |
| **SOUTH KOREA KSR** | Acute Treatment: door to needle time | 35 | 25 | 5 | 30 | 5 | 40 | 25 | 5 | 30 | 0 | - | | | | EXCLUDE |
| **UK SSNAP** | Was the patient given anticoagulant reversal therapy? | 35 | 15 | 10 | 35 | 5 | 40 | 15 | 10 | 35 | 0 | - | | | | EXCLUDE |
| **SOUTH KOREA CRCS-5** | Study ID | 10 | 40 | 5 | 35 | 10 | 5 | 50 | 5 | 35 | 5 | - | | | | EXCLUDE |

| | | | | | | | | | | | | | | | | |
|---|---|---|---|---|---|---|---|---|---|---|---|---|---|---|---|---|
| **SOUTH KOREA KSR** | Stroke subtype: Haemorrhage-ICH | 50 | 20 | 5 | 20 | 5 | 55 | 20 | 5 | 20 | 0 | 35 | 41 | 24 | 0 | EXCLUDE |
| **SOUTH KOREA KSR** | Stroke subtype: Haemorrhage-SAH | 50 | 20 | 5 | 20 | 5 | 55 | 20 | 5 | 20 | 0 | 35 | 41 | 24 | 0 | EXCLUDE |
| **SOUTH KOREA KSR** | Stroke subtype: TIA | 45 | 30 | 0 | 20 | 5 | 50 | 30 | 0 | 20 | 0 | 41 | 41 | 18 | 0 | EXCLUDE |
| **SOUTH KOREA KSR** | Acute Treatment: other reasons for no IV tPA - text | 10 | 30 | 20 | 35 | 5 | 20 | 25 | 20 | 35 | 0 | - | | | | EXCLUDE |
| **SOUTH KOREA KSR** | Vital/physical: height | 20 | 45 | 5 | 25 | 5 | 15 | 55 | 5 | 25 | 0 | - | | | | EXCLUDE |
| **SOUTH KOREA KSR** | Laboratory test: Hb | 10 | 45 | 5 | 35 | 5 | 5 | 50 | 10 | 35 | 0 | - | | | | EXCLUDE |
| **SOUTH KOREA KSR** | Laboratory test: Total Chol | 25 | 35 | 10 | 25 | 5 | 20 | 40 | 15 | 25 | 0 | - | | | | EXCLUDE |
| **SOUTH KOREA KSR** | Laboratory test: HDL Chol | 20 | 30 | 15 | 30 | 5 | 15 | 35 | 20 | 30 | 0 | - | | | | EXCLUDE |
| **SOUTH KOREA KSR** | Laboratory test: TG | 15 | 40 | 10 | 30 | 5 | 15 | 45 | 10 | 30 | 0 | - | | | | EXCLUDE |
| **SOUTH KOREA KSR** | Laboratory test: LDL Chol | 20 | 35 | 10 | 30 | 5 | 15 | 40 | 15 | 30 | 0 | - | | | | EXCLUDE |
| **SOUTH KOREA KSR** | Laboratory test: fasting glucose | 10 | 35 | 20 | 30 | 5 | 10 | 40 | 20 | 30 | 0 | - | | | | EXCLUDE |
| **ONTARIO OSR** | Lab Investigations (subsequent tests to be considered following patient arrival to ED): HbA1C | 15 | 30 | 20 | 30 | 5 | 10 | 35 | 25 | 30 | 0 | - | | | | EXCLUDE |
| **SOUTH KOREA KSR** | Laboratory test: Cr | 15 | 40 | 10 | 30 | 5 | 5 | 45 | 20 | 30 | 0 | - | | | | EXCLUDE |
| **SOUTH KOREA CRCS-5** | PT (Similar items aPTT & INR) | 10 | 40 | 10 | 35 | 5 | 10 | 45 | 10 | 35 | 0 | - | | | | EXCLUDE |
| **SCOTLAND SSCA** | Demographics: Ethnicity | 55 | 15 | 10 | 15 | 5 | 60 | 15 | 10 | 15 | 0 | 59 | 18 | 23 | 0 | EXCLUDE |
| **SCOTLAND SSCA** | Patient Pathway: Did the patient have a carotid intervention? | 30 | 35 | 0 | 30 | 5 | 40 | 30 | 0 | 30 | 0 | - | | | | EXCLUDE |
| **SCOTLAND SSCA** | Carotid Intervention: Date of carotid intervention | 35 | 30 | 0 | 30 | 5 | 35 | 30 | 0 | 35 | 0 | - | | | | EXCLUDE |
| **USA GWTG** | First Glasgow Coma Scale (GCS) | 20 | 40 | 15 | 20 | 5 | 30 | 35 | 15 | 20 | 0 | - | | | | EXCLUDE |
| **USA PCNASP** | Complications of thrombolytic therapy: Were there bleeding complications in a patient transferred after IV tPA | 50 | 20 | 5 | 20 | 5 | 55 | 20 | 5 | 20 | 0 | 41 | 35 | 24 | 0 | EXCLUDE |
| **-AUSTRIA ASUR** | ASPECT score | 45 | 20 | 5 | 25 | 5 | 45 | 25 | 5 | 25 | 0 | - | | | | EXCLUDE |

| | | | | | | | | |
|---|---|---|---|---|---|---|---|---|
| ADDITIONAL EXPERT-SUGGESTED | Was this an emergency admission for stroke? | - | - | 53 | 18 | 23 | 6 | EXCLUDE |
| ADDITIONAL EXPERT-SUGGESTED | Was this a FAST-positive ambulance admission or walk in or other emergency that later turned out to be stroke? | - | - | 65 | 12 | 23 | 0 | EXCLUDE |
| ADDITIONAL EXPERT-SUGGESTED | Was the stroke onset witnessed? | - | - | 65 | 17.5 | 17.5 | 0 | EXCLUDE |
| ADDITIONAL EXPERT-SUGGESTED | Was there delayed ambulance pick up? | - | - | 59 | 23 | 18 | 0 | EXCLUDE |
| ADDITIONAL EXPERT-SUGGESTED | Occupation | - | - | 35 | 41 | 24 | 0 | EXCLUDE |
| ADDITIONAL EXPERT-SUGGESTED | Level of education | - | - | 47 | 35 | 18 | 0 | EXCLUDE |
| ADDITIONAL EXPERT-SUGGESTED | Socioeconomic status | - | - | 29.5 | 41 | 29.5 | 0 | EXCLUDE |
| ADDITIONAL EXPERT-SUGGESTED | Date and time that the patient was found | - | - | 53 | 23 | 18 | 6 | EXCLUDE |
| ADDITIONAL EXPERT-SUGGESTED | Location of Intracerebral haematoma in ICH. | - | - | **70** | 12 | 18 | 0 | INCLUDE |
| ADDITIONAL EXPERT-SUGGESTED | Was imaging performed elsewhere first? | - | - | 35 | 35 | 30 | 0 | EXCLUDE |
| ADDITIONAL EXPERT-SUGGESTED | Was the patient transferred out for thrombectomy? | - | - | 59 | 12 | 29 | 0 | EXCLUDE |
| ADDITIONAL EXPERT-SUGGESTED | Was carotid revascularisation done within two weeks of symptom onset? | - | - | 47 | 24 | 29 | 0 | EXCLUDE |
| ADDITIONAL EXPERT-SUGGESTED | Did neurosurgery take place? If yes, what surgery? | - | - | 53 | 29 | 18 | 0 | EXCLUDE |
| ADDITIONAL EXPERT-SUGGESTED | Was the patient catheterised? | - | - | 47 | 24 | 29 | 0 | EXCLUDE |

| | | | | | | | | |
|---|---|---|---|---|---|---|---|---|
| ADDITIONAL EXPERT-SUGGESTED | NIHSS at 24 hours, 5 days, discharge | - | - | 65 | 17 | 12 | 6 | EXCLUDE |
| ADDITIONAL EXPERT-SUGGESTED | mRS at 90 days | - | - | **82** | 18 | 0 | 0 | INCLUDE |
| ADDITIONAL EXPERT-SUGGESTED | Was screening for cognitive impairment completed using a valid screening measure? | - | - | **70** | 18 | 12 | 0 | INCLUDE |
| ADDITIONAL EXPERT-SUGGESTED | What cognitive screening tool was used? | - | - | 59 | 35 | 6 | 0 | EXCLUDE |
| ADDITIONAL EXPERT-SUGGESTED | What was the result/score of the cognitive assessment? | - | - | 65 | 29 | 6 | 0 | EXCLUDE |
| ADDITIONAL EXPERT-SUGGESTED | Was screening for aphasia conducted? | - | - | **70** | 18 | 12 | 0 | INCLUDE |
| ADDITIONAL EXPERT-SUGGESTED | Does the patient have aphasia? | - | - | **88** | 6 | 6 | 0 | INCLUDE |
| ADDITIONAL EXPERT-SUGGESTED | Dementia (diagnosis of pre-stroke dementia) | - | - | **76** | 12 | 12 | 0 | INCLUDE |

Note. The emboldened numbers represent those items that received ≥70% of the votes to Include as part of INAS. The numbers in red font represent those items for which there were vote changes made from Round 1 to Round 2. A reduced set of items were put forward for review in Round 3.

[a] Twenty out of 24 eligible stakeholders responded with item votes in the first round of the Delphi Consultation, representing an 83% response rate. The percentages represent the percentage of the 20 responding stakeholders who voted for each response option.

[b] Eleven out of the initially responding 20 stakeholders (55% response rate) responded with changed responses during the second round of the Delphi consultation. On average, 4.91% of the responses were changed by responding stakeholders; that is, they changed their vote from one eligible response to another. An additional 6.55% of responses were changed from an originally ‘missing’ vote to one of the eligible response options. Eight stakeholders did not respond, interpreted as no change to their previous responses. One additional stakeholder left their post after Round 1 and so their previous responses remained unchanged. The percentages displayed in the table represent the combined percentage votes across Rounds 1 and 2 for the 20 responding stakeholders including the initial votes for the 9 stakeholders who made no changes and the amended votes for the 11 responding participants in Round 2.
[c] The percentages represent the Delphi Round 3 votes for a reduced set of items and response options from 17 responding participants out of 22 eligible stakeholders (77% response rate).

Abbreviations: aPTT: activated partial thromboplastin clotting time; ASPECT: The Alberta Stroke Program Early CT Score; BP: blood pressure; Chol: cholesterol; Cr: creatinine; DBP: diastolic blood pressure; DVT: deep vein thrombosis; ECG: electrocardiogram; ED: emergency department; Hb: haemoglobin; HDL: high-density lipoprotein; IA: intra-arterial; ICH: intracerebral haemorrhage; INR: international normalised ratio; IV: intravenous; LPL: lipoprotein lipase; mRS: modified Rankin Scale; NIHSS: National Institutes of Health Stroke Scale; PT: prothrombin time; SAH: subarachnoid haemorrhage; SBP: systolic blood pressure; TG: triglycerides; TIA: transient ischaemic attack; TOAST: Trial of ORG 10172 in Acute Stroke Treatment; tPA: tissue plasminogen activator.

**Supplemental File H:** Prisma Scoping Review Checklist

### Preferred Reporting Items for Systematic reviews and Meta-Analyses extension for Scoping Reviews (PRISMA-ScR) Checklist

| SECTION | ITEM | PRISMA-ScR CHECKLIST ITEM | REPORTED ON PAGE # |
|---|---|---|---|
| **TITLE** | | | |
| Title | 1 | Identify the report as a scoping review. | Pg1 |
| **ABSTRACT** | | | |
| Structured summary | 2 | Provide a structured summary that includes (as applicable): background, objectives, eligibility criteria, sources of evidence, charting methods, results, and conclusions that relate to the review questions and objectives. | Pg5 |
| **INTRODUCTION** | | | |
| Rationale | 3 | Describe the rationale for the review in the context of what is already known. Explain why the review questions/objectives lend themselves to a scoping review approach. | Pgs 6-7 |
| Objectives | 4 | Provide an explicit statement of the questions and objectives being addressed with reference to their key elements (e.g., population or participants, concepts, and context) or other relevant key elements used to conceptualize the review questions and/or objectives. | Pgs 7-8 |
| **METHODS** | | | |
| Protocol and registration | 5 | Indicate whether a review protocol exists; state if and where it can be accessed (e.g., a Web address); and if available, provide registration information, including the registration number. | Pg 8 |
| Eligibility criteria | 6 | Specify characteristics of the sources of evidence used as eligibility criteria (e.g., years considered, language, and publication status), and provide a rationale. | Pg 8 |
| Information sources* | 7 | Describe all information sources in the search (e.g., databases with dates of coverage and contact with authors to identify additional sources), as well as the date the most recent search was executed. | Pgs 8-9 |
| Search | 8 | Present the full electronic search strategy for at least 1 database, including any limits used, such that it could be repeated. | Pg 9; Suppl. File A |
| Selection of sources of evidence† | 9 | State the process for selecting sources of evidence (i.e., screening and eligibility) included in the scoping review. | Pg 9 |
| Data charting process‡ | 10 | Describe the methods of charting data from the included sources of evidence (e.g., calibrated forms or forms that have been tested by the team before their use, and whether data charting was done independently or in duplicate) and any processes for obtaining and confirming data from investigators. | Pgs 9-10 |
| Data items | 11 | List and define all variables for which data were sought and any assumptions and simplifications made. | Pg 10 |

| SECTION | ITEM | PRISMA-ScR CHECKLIST ITEM | REPORTED ON PAGE # |
|---|---|---|---|
| Critical appraisal of individual sources of evidence§ | 12 | If done, provide a rationale for conducting a critical appraisal of included sources of evidence; describe the methods used and how this information was used in any data synthesis (if appropriate). | Pg 10 |
| Synthesis of results | 13 | Describe the methods of handling and summarizing the data that were charted. | Pgs 10-12; Suppl. File B |
| **RESULTS** | | | |
| Selection of sources of evidence | 14 | Give numbers of sources of evidence screened, assessed for eligibility, and included in the review, with reasons for exclusions at each stage, ideally using a flow diagram. | Pgs 13-14 |
| Characteristics of sources of evidence | 15 | For each source of evidence, present characteristics for which data were charted and provide the citations. | Pg 14 |
| Critical appraisal within sources of evidence | 16 | If done, present data on critical appraisal of included sources of evidence (see item 12). | N/A |
| Results of individual sources of evidence | 17 | For each included source of evidence, present the relevant data that were charted that relate to the review questions and objectives. | Pgs 14-15; Suppl. File C |
| Synthesis of results | 18 | Summarize and/or present the charting results as they relate to the review questions and objectives. | Pgs 15-17; Table 1, Figure 1 Suppl. Files D-G |
| **DISCUSSION** | | | |
| Summary of evidence | 19 | Summarize the main results (including an overview of concepts, themes, and types of evidence available), link to the review questions and objectives, and consider the relevance to key groups. | Pgs 18-20 |
| Limitations | 20 | Discuss the limitations of the scoping review process. | Pgs 20-21 |
| Conclusions | 21 | Provide a general interpretation of the results with respect to the review questions and objectives, as well as potential implications and/or next steps. | Pgs 21-22 |
| **FUNDING** | | | |
| Funding | 22 | Describe sources of funding for the included sources of evidence, as well as sources of funding for the scoping review. Describe the role of the funders of the scoping review. | Pg 23 |

JBI = Joanna Briggs Institute; PRISMA-ScR = Preferred Reporting Items for Systematic reviews and Meta-Analyses extension for Scoping Reviews.
* Where *sources of evidence* (see second footnote) are compiled from, such as bibliographic databases, social media platforms, and Web sites.
† A more inclusive/heterogeneous term used to account for the different types of evidence or data sources (e.g., quantitative and/or qualitative research, expert opinion, and policy documents) that may be eligible in a scoping review as opposed to only studies. This is not to be confused with *information sources* (see first footnote).
‡ The frameworks by Arksey and O'Malley (6) and Levac and colleagues (7) and the JBI guidance (4, 5) refer to the process of data extraction in a scoping review as data charting.
§ The process of systematically examining research evidence to assess its validity, results, and relevance before using it to inform a decision. This term is used for items 12 and 19 instead of "risk of bias" (which is more applicable to systematic reviews of interventions) to include and acknowledge the various sources of evidence that may be used in a scoping review (e.g., quantitative and/or qualitative research, expert opinion, and policy document).
